# Quantifying brain atrophy in Frontotemporal Dementia: a head-to-head comparison of neuroimaging techniques

**DOI:** 10.1101/2025.10.28.25339007

**Authors:** Amelie Metz, Roqaie Moqadam, Yashar Zeighami, D. Louis Collins, Sylvia Villeneuve, Mahsa Dadar

## Abstract

Frontotemporal Dementia (FTD) is a neurodegenerative disorder characterized by extensive atrophy in the frontal and temporal lobes of the brain as well as high cerebrovascular burden. While anatomical Magnetic Resonance Imaging (MRI) is well established for quantifying brain atrophy in FTD, the variability in (pre-)processing methods limits the generalizability and comparability of findings. This study systematically compared the robustness and sensitivity of multiple widely used neuroimaging metrics, namely Deformation-Based Morphometry (DBM), Voxel-Based Morphometry (VBM), Cortical Thickness (CT), and segmentation-based grey matter Volumes, in detecting atrophy across FTD subtypes. We processed 732 T1-weighted MRI scans from 156 participants with FTD and 139 healthy controls from the Frontotemporal Lobar Degeneration Neuroimaging Initiative using our in-house pipeline PELICAN (Dadar et al., 2025) for volumetric measures and FreeSurfer version 7 (Fischl, 2012) for CT and grey matter segmentations. Visual quality control using consistent quality control images at each step of the pipelines revealed significantly higher failure rates for CT (38.52%) and FreeSurfer segmentations (23.63%) relative to PELICAN’s volumetric measures (2.04% DBM, 3.05% VBM). Failure rates differed between FTD subtypes and were related to pathological burden. Particularly for FreeSurfer, errors occurred predominantly in regions with high prevalence of atrophy and White Matter Hyperintensities. In PELICAN, the addition of a FTD-specific template as an intermediate step during nonlinear registration decreased the failure rates in this step in the FTD population. We then applied linear regression models to assess each metric’s sensitivity in detecting cross-sectional differences between FTD groups controls as well as linear mixed-effects models to determine which method is most sensitive to longitudinal anatomical changes. While CT yielded effect sizes comparable to VBM and DBM when analyzing the same subset of successfully processed scans, VBM and DBM demonstrated enhanced power to detect effects due to lower failure rates and higher participant retention in the full sample. Overall, we demonstrate that image processing methodology and pipeline selection profoundly influences effect sizes and statistical power to detect meaningful between-group differences or longitudinal changes. Volumetric measures (DBM and VBM) yielded sufficiently robust pipeline outcomes to maintain adequate statistical power for capturing atrophy patterns after quality control procedures.

## 1. Introduction

Frontotemporal dementia (FTD) is a group of clinical syndromes characterized by progressive impairments in language and/or abnormal changes in behavior (Bang et al., 2015; Olney et al., 2017). This umbrella term encompasses behavioral-variant FTD (bvFTD) and two forms of primary progressive aphasia (PPA): semantic-variant PPA (svPPA) and non-fluent variant PPA (nfvPPA) (Gorno-Tempini et al., 2011; Rascovsky et al., 2011). FTD is characterized by extensive neurodegeneration in the frontal and temporal lobes of the brain (Mackenzie et al., 2009) as well as neuropathological abnormalities in the form of hyperphosphorylated protein accumulations, typically composed of tau or TDP-43 (Mackenzie et al., 2010; Rademakers et al., 2012). Patients also exhibit increased levels of cerebrovascular pathologies, including leukoaraiosis which manifests as White Matter Hyperintensities (WMHs) in Fluid-Attenuated Inversion Recovery (FLAIR) or T2-weighted MRI scans (Dadar, Mahmoud, et al., 2022). While the value of anatomical MRI for both diagnosis and study of FTD has been well established (Meeter et al., 2017), there is considerable variability in the methods applied to (pre-)process and analyze MRI scans in the literature, limiting the generalizability and comparability of findings.

Several neuroimaging modalities and morphometric analysis techniques have been employed to quantify brain atrophy in FTD, including volume-based measures based on grey matter segmentation (Fischl et al., 2002), voxel-based morphometry (VBM) (Ashburner & Friston, 2000), and deformation-based morphometry (DBM) (Ashburner et al., 1998), and surface-based measures such as cortical thickness (CT) (Dale et al., 1999; Fischl et al., 1999).

### Deformation-based morphometry (DBM)

The principle of DBM is to warp each individual scan to a common template through linear and non-linear deformation following pre-processing of the native scan, where local shape differences between the two images (i.e., the participant’s T1-weighted (T1w) image and the template) are captured in the deformation fields. The local deformation obtained from the non-linear transformations can then be used as a measure of tissue expansion or contraction by estimating the determinant of the Jacobian for each transform (Ashburner et al., 1998). Local contractions can be interpreted as shrinkage of tissue (atrophy), and local expansions are often related to tissue growth, or ventricular or sulcal enlargement (Chung et al., 2001). DBM provides signal for cortical and subcortical grey matter as well as white matter. As DBM only relies on precise registration and typically does not involve smoothing, it can be sensitive enough to detect subtle systematic structural differences (Cardenas et al., 2007). On the other hand, the absence of spatial smoothing means that registration errors tend to have a stronger impact in DBM than in VBM, as smoothing helps mitigate small errors (Ashburner & Friston, n.d.).

### Voxel-based morphometry (VBM)

VBM is another widely used neuroimaging technique for voxel-wise estimation of regional tissue volumes (Ashburner & Friston, 2000). Same as DBM, VBM can be applied to cortical and subcortical grey matter as well as white matter. The standard workflow for VBM involves spatial normalization, tissue segmentation, and spatial smoothing, meaning VBM constitutes a combination of DBM followed by tissue segmentation and smoothing. To create DBM maps, individual scans are spatially normalized to a common stereotactic space using linear and nonlinear registration to a standard template. This step ensures voxel-wise comparability and corrects for global head size and shape-related differences. Next, tissue segmentation categorizes tissue into grey matter, white matter, and cerebrospinal fluid based on intensity values and derives tissue probability maps. Tissue probability maps are then nonlinearly transformed to the standard template space and modulated by the DBM maps to identify local atrophy or expansion in the individual scans. Lastly, the resulting maps undergo spatial smoothing using a Gaussian kernel. This is necessary to account for residual small-scale interindividual anatomical differences after spatial normalization and to ensure a normal distribution of residuals, enhancing the validity of parametric statistical tests (Ashburner & Friston, 2000; Kurth et al., 2015; Whitwell, 2009). The primary limitation of VBM is its reliance on tissue segmentation, whereby any tissue misclassification can lead to inaccurate VBM estimations (Dadar, Potvin, et al., 2021). Due to the increased prevalence of lesions WMHs in individuals with neurodegenerative disorders, these errors might occur more frequently in these populations, introducing a systematic bias in derived metrics (Dadar, Potvin, et al., 2021). Furthermore, subtle signals might be removed during spatial smoothing, making VBM less sensitive to small volumetric changes (Ashburner & Friston, 2000; Chung et al., 2001). Lastly, despite it often being referred to as a metric of tissue density, the biological interpretation of VBM is unclear and should not be confused with cell density assessed cytoarchitectonically, as VBM is computed as the product of tissue classification probability and local volume change (Mechelli et al., 2005; Schwarz & Kašpárek, 2011).

### FreeSurfer Cortical Thickness (CT) and Segmentation

CT is defined as the distance between the inside and outside cortical surfaces, i.e. the boundary between white matter and grey matter and the grey matter/pial surface boundary (Lerch, 2015). The most common method of mapping CT uses surface-based techniques whereby preprocessing steps include linear registration to stereotactic space, nonuniformity correction, and tissue classification (Lerch, 2015). CT is then estimated by creating two polyhedral surfaces along the inside and outside cortical boundaries and calculating the distance between these two surfaces at each vertex. Postprocessing involves nonlinear surface-based alignment, parcellation of the cortex, and surface-based smoothing of the thickness maps along the surfaces (Dale et al., 1999; Fischl, 2012; Fischl et al., 1999). For longitudinal data, FreeSurfer first performs initial preprocessing on each time point and then builds a within-subject template that represents the subject’s average anatomy. All preprocessing steps are then repeated for each scan, using this template as a common reference, to improve robustness and reduce bias (Reuter et al., 2012).

This method allows for easy vertex-wise comparison of CT across populations, which might be more anatomically meaningful than comparing intensity values. The main downsides are the computational complexity (Scanlon et al., 2011) and, similar to VBM, its dependency on initial tissue classification, so that tissue misclassifications can lead to downstream errors. Additionally, CT does not provide measurements of subcortical or white matter atrophy. Given that evidence suggests subcortical atrophy often precedes cortical degeneration in FTD (Planche et al., 2023) and may serve as an important imaging biomarker of disease progression (Manera et al., 2022), the exclusion of subcortical measures represents a significant limitation.

In addition to CT metrics, FreeSurfer also explicitly segments and provides volumetric measures for cortical and subcortical grey matter structures. Following the aforementioned preprocessing steps, FreeSurfer combines voxel intensity information with spatial priors to assign each voxel a neuroanatomical label using a Bayesian classification framework. Spatial priors are derived from a probabilistic atlas constructed from manually labeled training data (Fischl et al., 2002). This is a significant limitation given that the probabilistic atlas was trained on relatively small, healthy samples, potentially reducing the accuracy in populations with atrophy, lesions (Dadar, Potvin, et al., 2021), or atypical anatomy (King et al., 2020). Consequently, FreeSurfer segmentations need to undergo careful and time-intensive quality control (Vahermaa et al., 2023). Previous MRI studies on FTD have primarily focused on assessing brain atrophy through CT and VBM, demonstrating subtype-specific patterns of neurodegeneration (Chu et al., 2024; Meeter et al., 2017). BvFTD has been related to atrophy in frontal and insular cortices as well as the basal ganglia (Pan et al., 2012; Rosen et al., 2005; Schroeter et al., 2014; Seeley et al., 2008). SvPPA has been associated with left anterior temporal pole and hippocampal atrophy (Chan et al., 2001; Davies et al., 2009; Galton et al., 2001). In nfvPPA, atrophy seems to be most prevalent in the left inferior frontal gyrus, specifically involving Broca’s area (Akhmadullina et al., 2024; Bisenius et al., 2016; Mesulam et al., 2009; Rogalski et al., 2014). DBM has been applied in relatively few FTD studies, with results generally consistent with other MRI-based research (Cardenas et al., 2007; Manera et al., 2019; Shafiei et al., 2023; Wisse et al., 2021).

Given that VBM, DBM, and CT each have their own advantages and drawbacks, it remains unclear which of these three approaches would be best suited for studying FTD. While studies have compared the utility of different methods in other diseases, such as epilepsy (Scanlon et al., 2011) and multiple sclerosis (Righart et al., 2017), similar comparisons in the context of FTD are still lacking. Similarly, no systematic analysis of pipeline failure rates in FTD populations has been conducted. Considering that MRI processing workflows are typically optimized for healthy brains, they often struggle to accurately process scans with severe atrophy or lesions. This makes it essential to identify the method that can be most robustly applied to FTD individuals, who exhibit high levels of pathology, ensuring reliable and accurate imaging-derived measures in this population. These issues are especially important in clinical and longitudinal studies, where small participant samples lead to a lack of power to detect meaningful differences, and consistent detection of disease-related changes is essential for accurate diagnosis, patient stratification, and tracking disease progression.

To address these issues, we systematically compared the robustness and sensitivity of multiple widely used neuroimaging approaches (VBM, DBM, and FreeSurfer-derived CT and volume) in detecting atrophy across FTD subtypes. Using a cohort of individuals with clinically diagnosed FTD and cognitively unimpaired controls, we investigated which method results in the lowest pipeline failure rates while maintaining sufficient sensitivity to detect meaningful anatomical differences between FTD and healthy controls, as well as between FTD subtypes. Based on findings in other populations, we hypothesized that CT would be most sensitive in detecting distinctive patterns of cortical and subcortical atrophy in FTD and that DBM would achieve lower quality control failure rates than other techniques. Our goal was to provide an empirical foundation for selecting appropriate imaging tools in FTD research and clinical settings, and to highlight trade-offs between sensitivity and robustness that may influence methodological choices in future studies of neurodegenerative disease.

## 2. Materials and methods

### 2.1. Participants

The Frontotemporal Lobar Degeneration Neuroimaging Initiative (FTLDNI, also referred to as NIFD) was founded through the National Institute of Aging and started in 2010 (https://memory.ucsf.edu/research/studies/nifd). The primary goals of NIFD are to identify neuroimaging modalities and methods of analysis for tracking frontotemporal lobar degeneration and to assess the value of imaging versus other biomarkers in diagnostic roles. NIFD provides MRI as well as various clinical and cognitive data. The inclusion criteria for FTD patients were a diagnosis of possible or probable FTD according to the FTD consortium criteria (Gorno-Tempini et al., 2011; Rascovsky et al., 2011). Longitudinal data from 292 NIFD participants were included in this project. Data was accessed and downloaded through the LONI platform in July 2023. The cohort includes patients with bvFTD (n_baseline_ = 77), svPPA (n_baseline_ = 39), and nfvPPA (n_baseline_ = 40) as well as 136 healthy controls (see Table 1 for demographic and cognitive characteristics of this cohort). In total, 766 T1w MRI scans were analyzed.

**Table 1.** Baseline demographic and cognitive characteristics in FTD subtypes and healthy controls in the NIFD cohort. Values expressed as mean (standard deviation). Asterisks indicate significant group differences based on one-way ANOVA or χ2 analysis comparing the groups. bvFTD: behavioral-variant Frontotemporal Dementia, svPPA: semantic-variant Primary Progressive Aphasia, nfvPPA: nonfluent-variant Primary Progressive Aphasia.

|  |  | bvFTD | svPPA | nfvPPA | control | p-value |
| --- | --- | --- | --- | --- | --- | --- |
| **Number** |  | 77 | 39 | 40 | 139 |  |
| **Age (years)** |  | 61.3 (6.3) | 62.8 (6.1) | 67.6 (7.3) | 62.9 (7.3) | <0.001* |
| **N follow-up**<br>**(participants (scans))** |  | 60 (102) | 37 (71) | 29 (54) | 123 (244) |  |
| **Follow-up time (years)** |  | 1.30 (0.84) | 1.34 (0.47) | 1.37 (0.83) | 2.80 (2.20) | <0.001* |
| **Sex (Male(%)/Female(%))** |  | 51 (66.2) | 23 (59.0) | 18 (45.0) | 58 (42.6) | 0.006 |
|  |  | 26 (33.8) | 16 (41.0) | 22 (55.0) | 78 (57.4) |  |
| **Education (years)** |  | 15.6 (3.2) | 16.9 (3.1) | 16.5 (3.3) | 17.5 (1.9) | <0.001* |
| **Clinical Dementia**<br>**Rating** | **0** | 1 (1.3) | 0 (0.0) | 9 (22.5) | 88 (64.7) | <0.001* |
|  | **0.5** | 21 (27.3) | 28 (71.8) | 25 (62.5) | 5 (3.7) |  |
|  | **1** | 36 (46.8) | 9 (23.1) | 4 (10.0) | 0 |  |
|  | **2** | 17 (22.1) | 2 (5.1) | 1 (2.5) | 0 |  |

### 2.2. Acquisition of MR images

The FTLDNI uses the infrastructure established by the Alzheimer’s Disease Neuroimaging Initiative (ADNI). All participating imaging centers share a common platform. Available information on acquisition parameters and scanners is summarized in Supplementary Table 1. For further details on MRI acquisition protocols and scanner information, please refer to https://cind.ucsf.edu/research/grants/frontotemporal-lobar-degeneration-neuroimaging-initiative-0.

### 2.3. MRI processing

#### 2.3.1. PELICAN (VBM, DBM)

We utilized the Pipeline for Evaluating Longitudinal Images of Cerebral ANatomy (PELICAN) (Dadar et al., 2025), an open source extensively validated and widely used in-house pipeline for processing both cross-sectional and longitudinal imaging data (Fereshtehnejad et al., 2025; Kamal et al., 2025; Lajoie et al., 2025; Metz et al., 2024; Moqadam et al., 2024, 2025; Qiu et al., 2024). This pipeline is built on the open-source MNI MINC-Toolkit v2 (Neelin et al., 1998; Vincent et al., 2016) and ANTs (Avants et al., 2009) toolkits. Preprocessing steps include denoising with optimized non-local means filtering (Coupe et al., 2008), intensity inhomogeneity correction (Sled et al., 1998), and intensity normalization via linear histogram matching. Images are then linearly registered to the MNI152-2009c template (Fonov et al., 2011) (nine parameters: 3 translation, 3 rotation, and 3 scaling) using a hierarchical linear registration approach (Dadar, Fonov, et al., 2018). For longitudinal data, an individual-specific template is generated following these preprocessing steps. Subsequently, brain extraction from preprocessed T1w images is performed using the BEaST algorithm (Eskildsen et al., 2012). The extracted brains are then non-linearly registered to the MNI152-2009c template using ANTs nonlinear registration (Avants et al., 2009). DBM maps are derived by computing the determinant of the Jacobian deformation field at each voxel. Additionally, nonlinear registration is also performed with a specific FTD population template (Dadar, Manera, et al., 2021) as an intermediate registration target, and the final nonlinear transformation is calculated by concatenating the individual-to-indirect-template and indirect-template-to-average-template nonlinear transformations. This step was added to mitigate increased registration errors that can occur in populations with severe pathology when using average templates derived from healthy brains, such as the MNI152-2009c (Dadar, Fonov, et al., 2018; Dadar, Manera, et al., 2021; Dadar, Camicioli, et al., 2022). (Both direct and indirect registration pathways will be evaluated below.) Subsequently, the BISON pipeline (Dadar & Collins, 2021) is applied to the linearly registered images for tissue segmentation and WMH extraction. BISON enables robust tissue and WMH segmentation using either T1w images alone or a combination of T1w and either FLAIR or T2w images (Dadar, Maranzano, et al., 2018). Finally, VBM maps are created by multiplying the BISON tissue probability maps by the determinant of the Jacobian of the nonlinear deformation fields. To account for differences in head size between participants, similar to the approach used by FreeSurfer (Klasson et al., 2018), we estimated Total Intracranial Volume (TIV) by dividing the total number of voxels in the MNI152-2009c template mask (i.e. the template TIV) by the scaling factor used to linearly align a participant’s scans to the standard template.

*Scaling factor = x transform * y transform * z transform*

*Total Intracranial Volume (TIV) = total number of voxels in ICBM template mask (1,886,574 mm^3^)/scaling factor*

Visual quality control was performed on multiple steps of the processing workflow, including linear and nonlinear registration, brain mask extraction, and tissue segmentation. VBM and DBM measures were calculated both voxel-wise and based on the CerebrA atlas, which includes 102 cortical, subcortical, and white matter regions (Manera et al., 2020). As the CerebrA atlas was derived from the Desikan-Killiany-Tourville (DKT) atlas (Desikan et al., 2006), which is used by FreeSurfer, regional results from PELICAN are comparable with FreeSurfer regional outputs.

#### 2.3.2. FreeSurfer (CT, Volume)

Estimation of CT and grey matter volumes was performed using longitudinal FreeSurfer (Fischl, 2012) version 7.4.1. The preprocessing pipeline includes individual-specific template creation, brain extraction, linear registration, and intensity normalization. Following preprocessing, the gray/white matter boundary is tessellated, and automated topology correction is applied. The surface is then deformed along intensity gradients to accurately position the gray/white and gray/CSF borders, generating the cortical models (Dale et al., 1999; Fischl et al., 1999). CT, i.e., the distance between the two cortical boundaries, is then computed at the vertex level. We quality-controlled the reconstructed gray/white and gray/CSF borders, separately for anterior and posterior regions and left and right hemispheres, using *freeview* visualizations. Here, the borders were overlaid on the preprocessed T1w images, and the rater consistently scrolled through the slices in axial (top to bottom), coronal (anterior to posterior), and sagittal (right to left) views. Mismatches between the estimated and the actual gray/white or gray/CSF borders were noted overall, and separately for left/right and anterior/posterior regions. CT measures were extracted both vertex-wise and parcellated according to the Desikan-Killiany-Tourville atlas (Desikan et al., 2006).

Additionally, we analyzed FreeSurfer grey matter segmentations. Following pre-processing, the pipeline performs tissue segmentation using voxel intensities and tissue probabilities (Fischl et al., 2002). We visually quality-controlled linear registrations and tissue segmentations. Visual quality control was performed, similar to PELICAN, on linear registrations and tissue segmentations. Volumes were extracted based on the Desikan-Killiany-Tourville atlas (Desikan et al., 2006). To account for differences in head size between participants, we also extracted the estimated TIV value provided by FreeSurfer for each participant.

### 2.4. Statistical Analyses

#### 2.4.1. Robustness of processing pipelines

Pipeline outputs underwent visual quality control, and failure rates across the three methodologies were compared using chi-square tests. Demographic (age and sex) and clinical characteristics of participants whose scans passed versus failed quality control were compared using one-way ANOVA for continuous variables and chi-square tests for categorical variables. For clinical differences in the FTD cohorts, we assessed disease severity based on the sum of boxes score of the Clinical Dementia Rating. We also compared grey matter volumes and White Matter Hyperintensity burden, which was extracted from FLAIR images using PELICAN (BISON) (Dadar & Collins, 2021).

#### 2.4.2. Correlation between methods

To assess the concordance between morphometric estimates derived from the different image-processing approaches, we conducted correlation analyses between regional measures obtained from PELICAN and FreeSurfer for the baseline visit of each participant. To achieve this, we computed region-wise partial correlations. For each brain region and pair of imaging methods, we modeled the association between the two regional measures both for the entire dataset as well as separately for cognitively normal participants and FTD patients. Within each group, we fit two linear regression models of the form

*Measure(method A) ∼ Measure(method B) + Total Intracranial Volume (TIV)*

For all combinations between DBM (indirect registration), DBM (direct registration), VBM, CT, and segmentation-based volumes. All measures and TIV were z-scored. Residuals from these models, representing variation in each measure unexplained by total intracranial volume, were extracted and correlated using Spearman correlation. This yielded a partial Spearman correlation for each region and method pair within the complete sample and each diagnostic group. Region-wise correlations were subsequently aggregated using Fisher’s z-transformation to obtain group-level summary statistics. Furthermore, we evaluated the agreement between TIV estimates of PELICAN and FreeSurfer using Spearman correlation.

#### 2.4.3. Sensitivity to group differences

To assess the sensitivity of each method in detecting differences in brain atrophy between FTD and healthy participants, as well as across FTD subtypes, we quantified and compared the effect sizes of diagnostic group differences across methods. In the case of DBM, analyses were conducted separately for measures derived using direct registration to the average template and those obtained via indirect registration using a disease-specific template. The following linear regression models were applied to cross-sectional data, separately for each neuroimaging method, comparing atlas-based and voxel-wise/vertex-wise atrophy measures across different regions:

*Measure ∼ 1 + Diagnostic Group + Age + Sex + Total Intracranial Volume (TIV)*

where the term Measure indicated the z-scored atrophy values derived from each method at baseline visits. Diagnostic Group was the variable of interest, distinguishing healthy controls from FTD patients. Age, Sex, and TIV (Brzezinski-Rittner et al., 2025) were added as covariates. Results were corrected for multiple comparisons using False Discovery Rate (FDR)(Genovese et al., 2002) correction with a significance threshold of p ≤ 0.05.

To ensure valid comparisons between methods while accounting for how increasing sample size with more robust pipelines affects statistical power, we conducted the analysis in two ways. First, we included all participants and scans that met quality control standards for each respective method, and second, we included only participants and scans that passed visual quality control across all methods. Numbers of participants/scans included in each part of the analyses can be found in Supplementary Figure 2.

#### 2.4.4. Sensitivity to temporal change in brain atrophy

In order to assess the differences between the neuroimaging techniques in terms of detecting neurodegenerative changes in brain structure over time, the following linear mixed-effects models were applied, separately for each neuroimaging method, comparing atlas-based and voxel-wise atrophy measures across different regions:

*Measure ∼ 1 + Diagnostic Group : Time from Baseline + Diagnostic Group + Time from Baseline + Diagnostic Group : Age at Baseline + Age at Baseline + Sex + Total Intracranial Volume (TIV) + (1|Participant ID)*

where the term Measure indicated the z-scored values derived from each method at different participant timepoints. The interaction between Diagnostic Group and Time from Baseline (*Diagnostic Group : Time from Baseline*) was the term of interest, assessing the longitudinal slope in each FTD variant relative to healthy controls. Sex and TIV were added as covariates. Results were corrected for multiple comparisons using FDR (Benjamini & Hochberg, 1995; Genovese et al., 2002) correction with a significance threshold of p ≤ 0.05. Again, the analysis was performed both in all participants and in the subset of participants that met quality control standards across all methods. We also excluded follow-up visits that occurred later than 3.9 years after the baseline visit (90th percentile), to ensure a similar distribution of follow-up times between participants.

## 3. Results

### 3.1. Quality control failure rates

After visually assessing raw MRI scans and excluding scans with significant artifacts due to motion or incidental findings (e.g. stroke), a total of 732 scans remained for analysis. Run times varied considerably between the image processing methods. PELICAN processed participants with a single scan within 2 hours and required another 1.5-2 hours for each additional scan. FreeSurfer proved substantially more time-intensive, requiring up to 45 hours for a participant’s initial two timepoints and an additional 15 hours for each subsequent scan.

Examples of quality control images, both successful and failed, are included in Figure 1 and Supplementary Figure S4. Quality control pass and fail rates for VBM, DBM, CT, and FreeSurfer segmentations are shown in Table 2. Using PELICAN, all scans passed brain mask creation. Only 3 scans failed linear registration to the MNI template. However, 50 scans (6.83%) failed direct nonlinear alignment to the average template, whereas only 15 (2.05%) failed nonlinear registration using the indirect FTD-specific average template. Notably, the majority of failed scans in direct nonlinear registration were from FTD participants (80.77% of failures), while failures in indirect nonlinear registration mostly occurred in healthy controls (80%) (Figure 3). Additionally, 12 (1.64%) scans resulted in erroneous tissue segmentations. Overall, DBM processing failed in 50 cases (6.83%) using direct nonlinear registration and 15 cases (2.04%) using indirect nonlinear registration, while VBM processing failed in 62 cases (6.56%) for direct and 25 cases (3.05%) for indirect nonlinear registration. Chi-square analyses revealed significant differences in failure rates between the use of direct and indirect registration to the MNI template both for VBM (p<0.001) and DBM (p<0.001) but no significant difference in robustness between DBM and VBM (p=0.122).

**Figure 1.**
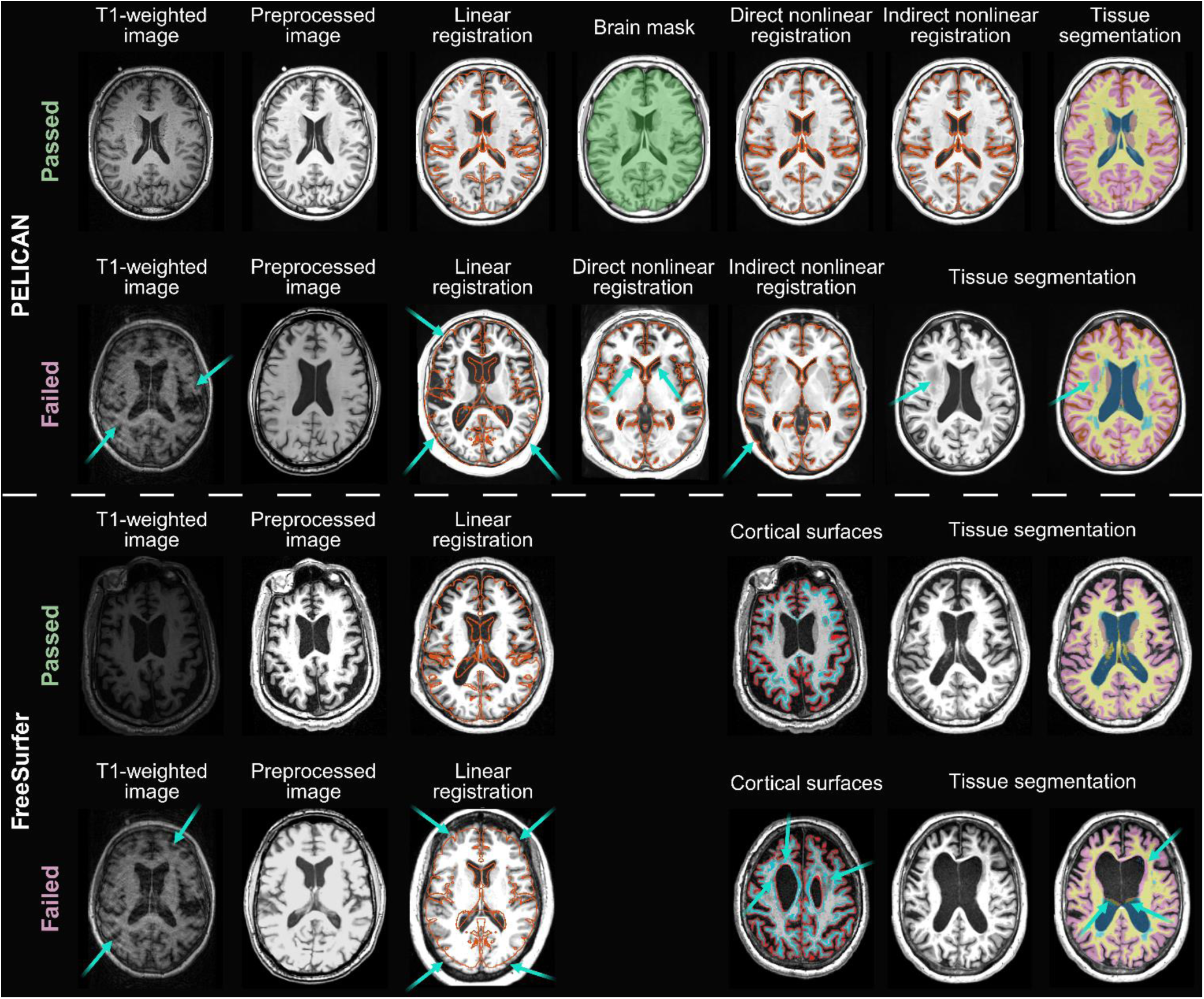
Examples of passed (left) and failed (right) quality control images at different steps of the pipelines. Cyan arrows indicate areas of failure. **PELICAN. Top row**, left to right: 1. Raw T1w image. 2. Pre-processed T1w image after denoising, intensity inhomogeneity correction, and intensity normalization. 3. Contours of the MNI-ICBM152 average template overlaid in red on the linearly registered image, showing accurate alignment. 4. BEaST brain mask overlaid on the T1w image in green. 5. Contours of the MNI-ICBM152 average template overlaid in red on the non-linearly registered image using direct registration to the MNI-ICBM space, showing accurate alignment. 6. Contours of the MNI-ICBM152 average template overlaid in red on the non-linearly registered image using indirect registration to the MNI-ICBM space through the NIFD-FTD average, showing accurate alignment. 7. BISON tissue segmentation map overlaid on the preprocessed image. **Bottom row**, left to right, scans from different participants: 1. Raw T1w image, showing high amounts of motion that interfere with processing pipelines. 2. Pre-processed T1w image after denoising, intensity inhomogeneity correction, and intensity normalization, showing a failure in intensity normalization. 3. Contours of the MNI-ICBM152 average template overlaid in red on the linearly registered image, showing inaccurate alignment (skull included within the template space). 4. Contours of the MNI-ICBM152 average template overlaid in red on the non-linearly registered image using indirect registration to the MNI-ICBM space, showing inaccurate alignment of the ventricles. 5. Contours of the MNI-ICBM152 average template overlaid in red on the non-linearly registered image using indirect registration to the MNI-ICBM space through the NIFD-FTD average, showing inaccurate alignment in the occipital/temporal lobes. 6. & 7. Preprocessed T1w image and BISON tissue segmentation map overlaid on the same image, showing errors in tissue segmentation whereby White Matter Hypointensities were labelled as grey matter. **FreeSurfer**. Similar quality control images for different FreeSurfer steps. The Cortical Thickness panel shows FreeSurfer inner and outer surfaces overlaid on the T1w image in native space. Note the inaccuracies in the failed image (bottom row) indicated by cyan arrows.

**Figure 2.**
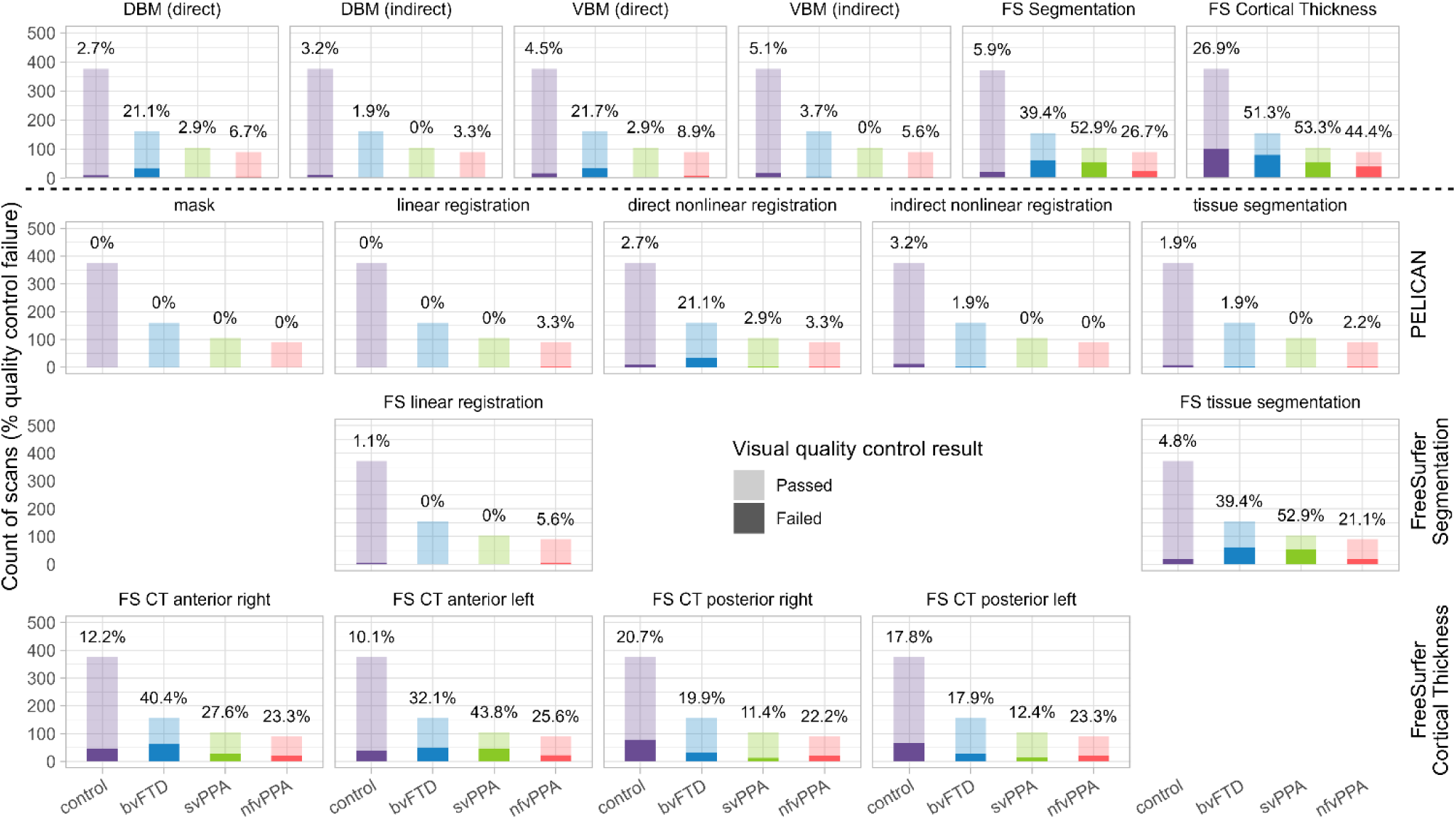
Barplots summarizing the results of quality control of MRI processing methods per diagnostic group. Transparent bars indicate the total number of T1w MRI scans per group, saturated bars show the number of scans that failed visual quality control. Percentages denote the portion of scans per group per processing step that failed quality control. Upper row showing full results for each method, lower three rows showing quality control results for different steps of each pipeline. VBM: Voxel-Based Morphometry, DBM: Deformation-Based Morphometry, CT: Cortical Thickness, FS: FreeSurfer, bvFTD: behavioral-variant Frontotemporal Dementia, svPPA: semantic-variant Primary Progressive Aphasia, nfvPPA: nonfluent-variant Primary Progressive Aphasia.

**Figure 3.**
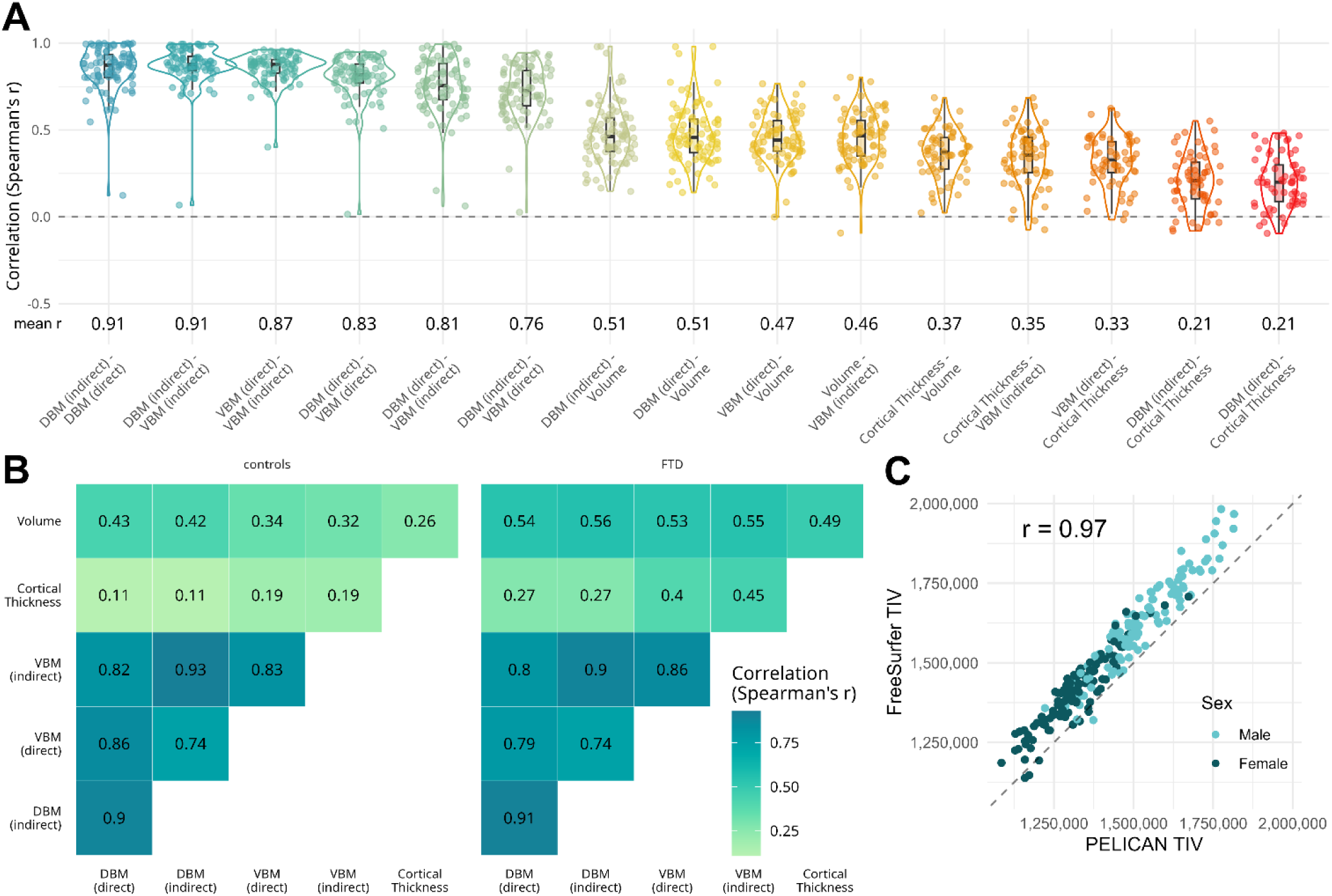
Correlations between metric derived from different neuroimaging methods. A) Distribution of region-wise correlations between morphometric measures derived from DBM (direct or indirect nonlinear registration), VBM, Cortical Thickness, and FreeSurfer Volumes. Each datapoint represents a pair-wise Pearson’s correlation r-value for estimates of two methods for one of the CerebrA/DKT atlas regions. Mean r values were calculated using Fisher’s z-transform. B) Mean Spearman’s correlation r-value between regional measures of the different methods based on Fisher z-transform, divided by healthy controls (left) and FTD participants (right). C) Comparison between estimated Total Intracranial Volumes (in mm^3^) derived from FreeSurfer and from PELICAN. Each datapoint represents one participant and colours denote each individual’s sex. VBM: Voxel-Based Morphometry, DBM: Deformation-Based Morphometry, CT: Cortical Thickness, TIV: Total Intracranial Volume.

**Table 2.**
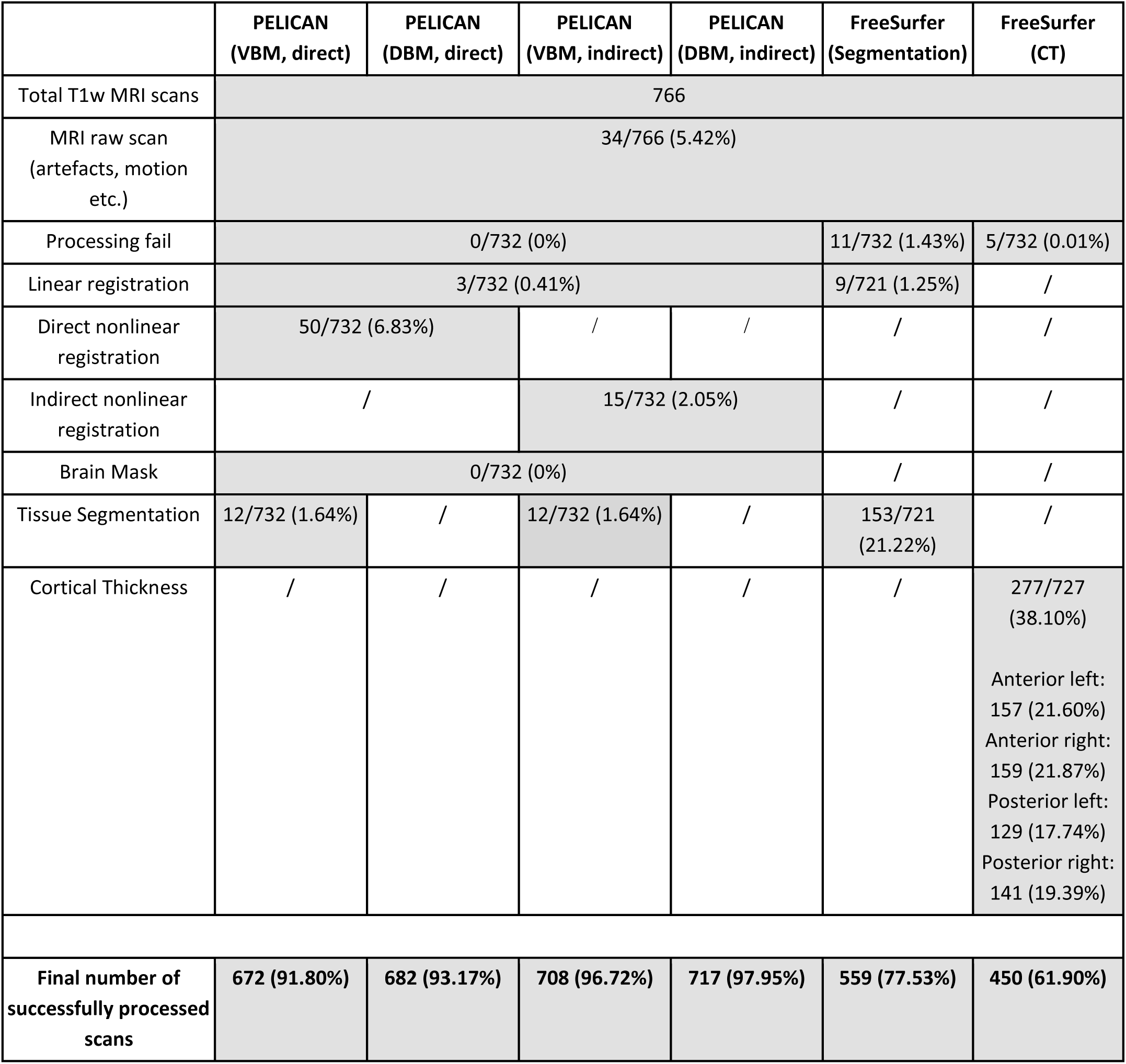
Results of quality control of MRI processing methods. Numbers and percentages of failed cases. VBM: Voxel-Based morphometry, DBM: Deformation-Based Morphometry, CT: Cortical Thickness, WMH: White Matter Hypointensity, BISON: Brain tissue segmentation pipeline (Dadar & Collins, 2021).

FreeSurfer failed to process 11 scans for segmentations and 5 for CT whereby the pipeline did not produce any outputs, despite multiple attempts to re-run. Among the scans that were fully processed, errors in linear registration occurred in 9 scans (1.25%) and tissue segmentation failed in 153 cases (21.22%). Quality control of CT maps showed misaligned grey matter/white matter or grey matter/pial surface estimations in 277 scans (38.10%) whereby 40-50% of patient scans were erroneous. Hence, FreeSurfer volume calculations failed for 173 scans (23.63%) and cortical thickness processing failed for 282 (38.52%) cases. Due to the high failure rate for FreeSurfer CT, we conducted regional quality control separately for anterior (frontal, temporal) and posterior (parietal, occipital) regions. The regional failure rates were: Anterior left: 157 scans (21.60%), anterior right: 159 scans (21.87%), posterior left: 129 scans (17.74%), posterior right: 141 scans (19.39%), whereby anterior regions showed more errors in patients, while posterior regions showed more errors in controls (Figure 3). Results of the chi-square analysis revealed that FreeSurfer volume processing yields lower failure rates than FreeSurfer CT (p<0.001). Furthermore, failure rates for DBM and VBM are significantly lower than either FreeSurfer volume or FreeSurfer CT (p<0.001).

When comparing demographic and clinical characteristics of participants whose scans passed versus failed visual quality control for either both or one of the pipelines, we found an association with disease severity as reflected by CDR sum of boxes, cerebrovascular burden, and sex (Table 3, Supplementary Figure S3), whereby scans from participants with higher Clinical Dementia Rating, higher WMH load, and male sex more frequently lead to processing errors.

**Table 3.** Demographic and clinical characteristics of participants whose scans failed in either one, both or neither of the tested neuroimaging pipelines FreeSurfer and PELICAN. Values expressed as mean (standard deviation). P-values are based on one-way ANOVA or χ2 analysis comparing the groups. Asterisks indicate significant group differences between the respective group and the “FreeSurfer & PELICAN Success” group based on t-tests.

|  | FreeSurfer & PELICAN Fail | FreeSurfer Fail | PELICAN Fail | FreeSurfer & PELICAN Success | p-value |
| --- | --- | --- | --- | --- | --- |
| Sex (Male:Female (%Female)) | 10:5 (33.3) | 65:42 (39.3) | 4:3 (42.9) | 60:81 (57.4) | 0.022 |
| Age (years) | 65.3 (6.3) | 64.8 (6.3) | 62.2 (10.8) | 63.0 (8.0) | 0.210 |
| Clinical Dementia Rating - Sum of Boxes (years) | 7.8 (4.2)* | 3.4 (3.4)* | 4.8 (6.2) | 1.9 (2.9) | <0.001 |
| Total White Matter Hyperintensity Volume (log-transformed) | 9.7 (0.8)* | 8.4 (1.2)* | 8.4 (1.8) | 7.8 (1.1) | <0.001 |
| Total Grey Matter Volume (mm<sup>3</sup>) | 1,565,750.3 (96,610.4) * | 1,477,231.4 (93,647.6)* | 1,471,788.6 (149,438.4) | 1,348,760.2 (105,468.6) | <0.001 |

### 3.2. Correlation between methods

We evaluated the inter-method agreement for regional metrics derived from DBM, VBM, CT, and segmentations based on the CerebrA/DKT atlas by calculating pair-wise partial correlations, adjusting for variation due to TIV (Figure 3A). The strongest correlations were observed among measures derived from PELICAN, whereby DBM using direct versus indirect registration as well as indirect DBM and VBM showed a mean correlation of r = 0.91, direct DBM versus VBM had a mean r = 0.83, and direct versus indirect VBM had a mean r = 0.87. In contrast, correlations between PELICAN- and FreeSurfer-derived volumetric measures (DBM/VBM versus FreeSurfer volumes) were lower, ranging from mean r = 0.46 to r = 0.51. Correlations between volume-based and surface-based metrics were generally low (mean r (Volume vs CT) = 0.37, mean r (VBM (direct) vs CT) = 0.35, mean r (VBM (indirect) vs CT) = 0.33). The lowest correlations overall were observed between DBM and CT (mean r = 0.21).

Figure 3B shows how the inter-method agreement differs among healthy controls and FTD patients. While we found that the patterns for PELICAN measure correspondence were overall similar between group, the mean correlation FreeSurfer outcomes were higher in the FTD group (e.g. Volume vs VBM (indirect): mean r (controls) = 0.32, mean r (FTD) = 0.55; CT vs VBM (indirect): mean r (controls) = 0.19, mean r (FTD) = 0.45).

As PELICAN and FreeSurfer both estimate TIV using the scaling factors during linear registration, we also compared their estimates (Figure 3C). While the correlation between both was high (r = 0.97), FreeSurfer estimates were consistently higher than PELICAN’s. This may result from differences in how FreeSurfer and PELICAN define intracranial space, or from the fact that FreeSurfer estimations are based on the MNI-305 template (https://surfer.nmr.mgh.harvard.edu/fswiki/eTIV), which is larger than MNI-ICBM152. Hence, FreeSurfer-based TIV estimates should not be compared to or combined with TIV estimates from other softwares.

### 3.3. Sensitivity to group differences

The sensitivity of each method for detecting differences between FTD variants and healthy controls was assessed using linear regression models. In the regional analysis of the matched subset of scans successfully processed by all methods, VBM yielded the largest effect sizes (e.g., direct VBM: range(bvFTD) = [-0.97,-0.03], range(svPPA) = [-2.84,0.28], range(nfvPPA) = [-1.39,-0.02]) (Figure 4A/B, for t-statistics see Supplementary Figures S5/S6) across all FTD groups, followed by CT (range(bvFTD) = [-1.14,0.06], range(svPPA) = [-4.41,0.22], range(nfvPPA) = [-0.77,0.23]). Accordingly, VBM identified the most regions as significantly different after FDR correction (e.g. direct VBM n(bvFTD) = 62, n(svPPA) = 29, n(nfvPPA) = 61, Figure 4C), followed by DBM (e.g., direct DBM: n(bvFTD) = 32, n(svPPA) = 26, n(nfvPPA) = 28) and CT (n(bvFTD) = 42, n(svPPA) = 15, n(nfvPPA) = 17).

**Figure 4.**
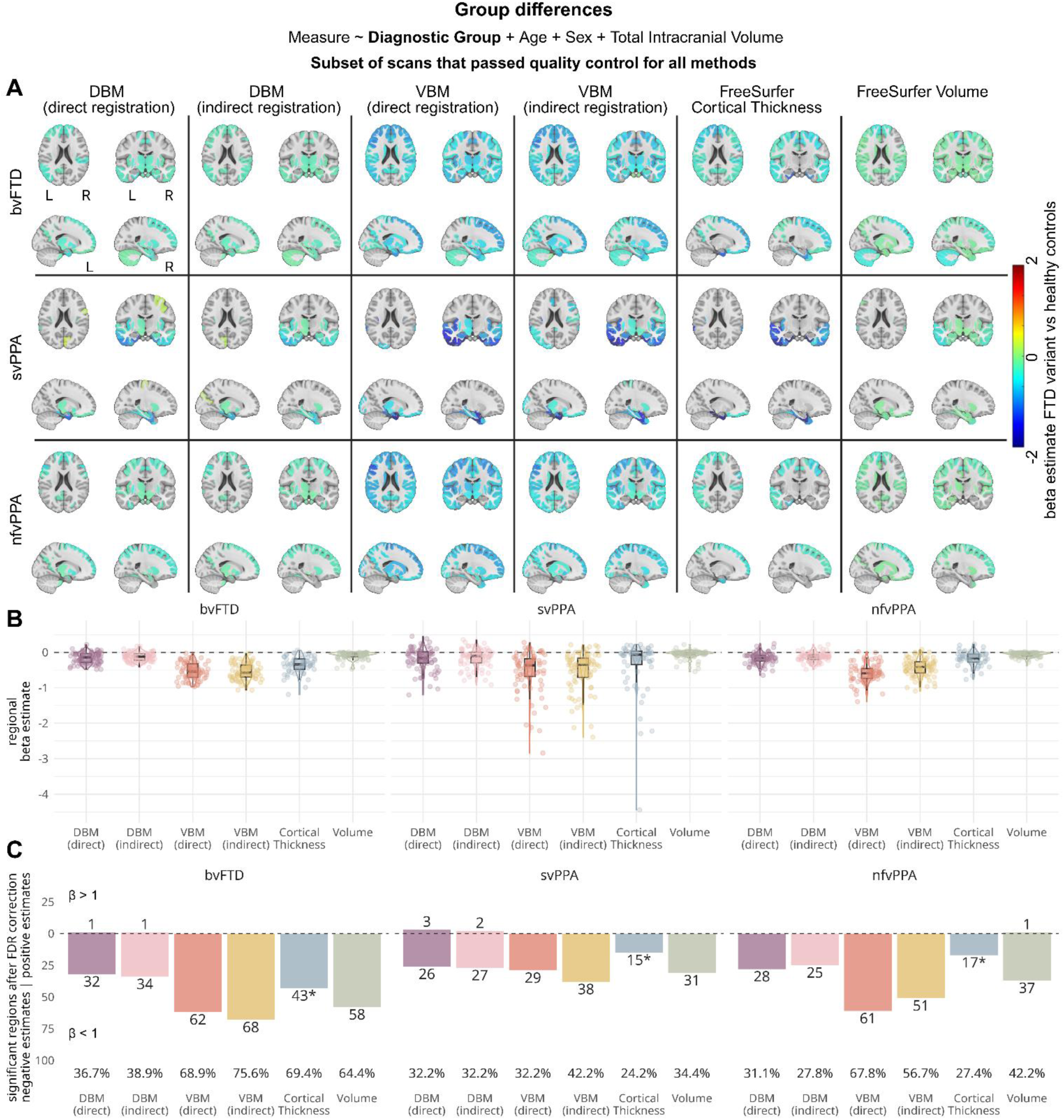
Results of linear regression models assessing the sensitivity of each method to detect differences between FTD subtypes and healthy controls in the subset of MRI scans that were successfully processed by each method. A) Brain maps showing beta estimates for the main effect for diagnostic group, comparing regional values for FTD variants versus healthy controls. B) Box- and violin plots summarizing beta estimates for the group effect for each method. Each datapoint represents the estimate for one atlas region. C) Barplots showing the number of regions with significant group differences, after FDR correction, divided by direction of the effect. Percentages of significant regions out of all atlas regions, regardless of effect direction, are shown below each bar. *Note that Cortical Thickness is only calculated for 62 of 90 regions, as this measure does not include subcortical areas. VBM: Voxel-Based Morphometry, DBM: Deformation-Based Morphometry, bvFTD: behavioral-variant Frontotemporal Dementia, svPPA: semantic-variant Primary Progressive Aphasia, nfvPPA: nonfluent-variant Primary Progressive Aphasia.

These patterns remained consistent in the full sample analysis (Figure 5A/B). However, the increased sample size and statistical power enabled VBM, DBM, and FreeSurfer Volumes to identify more significant regions, particularly in bvFTD and svPPA (Figure 5C), which had the highest failure rates and lowest numbers in the subset analysis. Sample size, and accordingly statistical power, for CT only marginally increased in the full sample analysis, so results for CT stayed widely consistent. In the voxel-/vertex-wise analysis, VBM and CT identified similar atrophy patterns across FTD variants, with predominant frontal lobe atrophy in bvFTD, temporal lobe atrophy in svPPA, and scattered frontotemporal involvement in nfvPPA (Figure 6A). Furthermore, VBM and CT detected significant differences across a larger number of voxels/vertices than DBM (e.g., subset analysis in bvFTD: percent(direct DBM) = 6.6%, percent(indirect DBM) = 7.1%, percent(direct VBM) = 17.3%, percent(indirect VBM) = 29.1%, percent (CT) = 32.6%, Supplementary Figure 7), although DBM showed improved power and VBM outperformed CT in the full sample, whereby indirect nonlinear registration provided improved results (e.g. bvFTD: percent(direct DBM) = 25.3%, percent(indirect DBM) = 31.6%, percent(direct VBM) = 61.5%, percent(indirect VBM) = 69.2%, percent (CT) = 32.6%, Figure 6C). VBM produced larger effect sizes than CT (e.g. subset analysis indirect VBM: range(bvFTD) = [-1.63,0.72], range(svPPA) = [-2.19,0.88], range(nfvPPA) = [-1.46,0.75]; subset analysis CT: range(bvFTD) = [-1.15, 1.08], range(svPPA) = [-2.53,0.72], range(nfvPPA) = [-1.07,1.27]), Figure 6B/Supplementary Figure 7, for t-statistics see Supplementary Figures S8/S9). DBM showed fewer significant voxelwise differences and notably identified effects in the opposite direction compared to other methods, particularly in periventricular regions adjacent to large sulci (see for instance hippocampus in svPPA).

**Figure 5.**
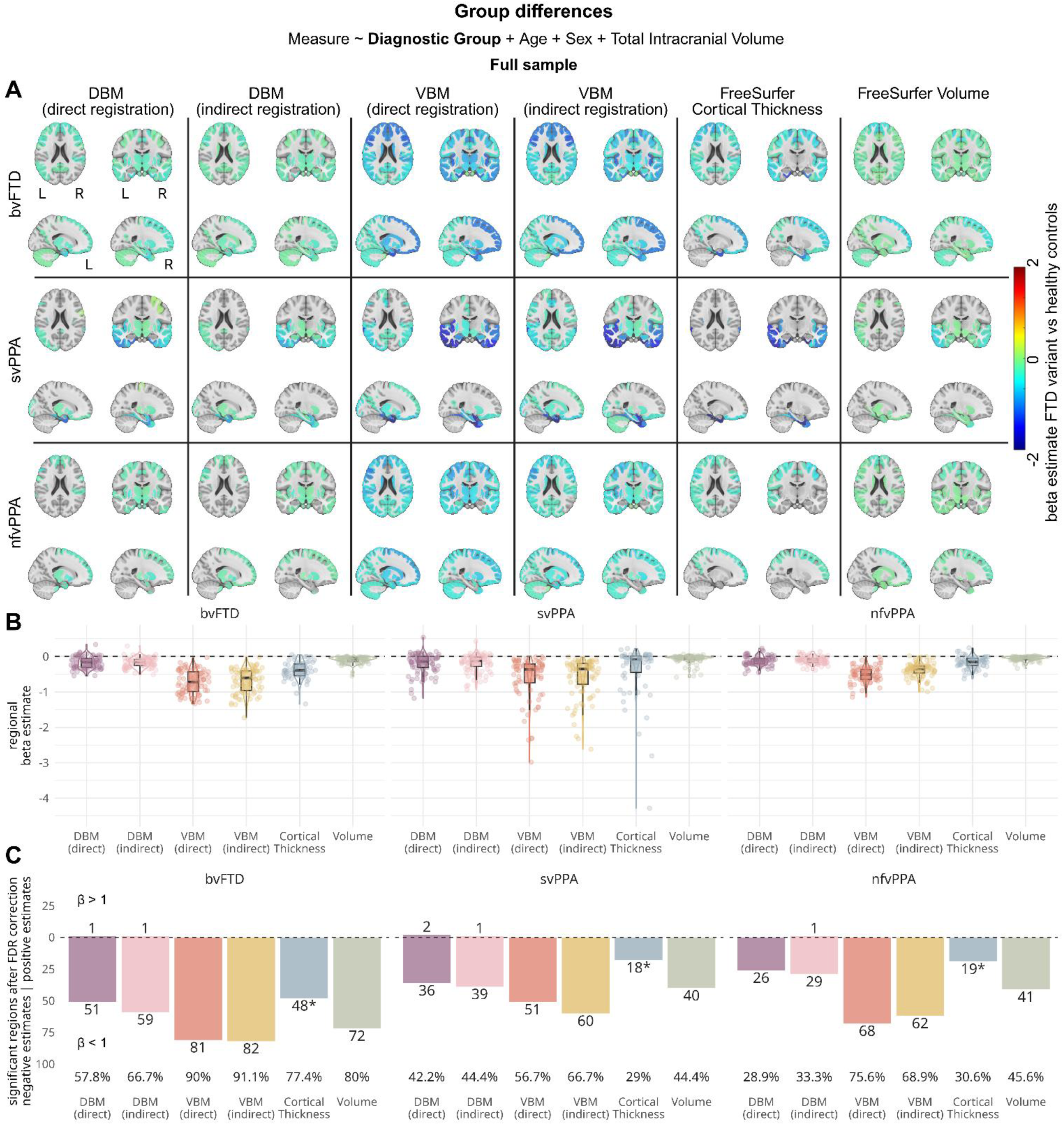
Results of linear regression models assessing the sensitivity of each method to detect differences between FTD subtypes and healthy controls in the full sample. A) Brain maps showing beta estimates for the main effect for diagnostic group, comparing regional values for FTD variants versus healthy controls. B) Box- and violin plots summarizing beta estimates for the group effect for each method. Each datapoint represents the estimate for one atlas region. C) Barplots showing the number of regions with significant group differences, after FDR correction, divided by direction of the effect. Percentages of significant regions out of all atlas regions, regardless of effect direction, are shown below each bar. *Note that Cortical Thickness is only calculated for 62 of 90 regions, as this measure does not include subcortical areas. VBM: Voxel-Based Morphometry, DBM: Deformation-Based Morphometry, bvFTD: behavioral-variant Frontotemporal Dementia, svPPA: semantic-variant Primary Progressive Aphasia, nfvPPA: nonfluent-variant Primary Progressive Aphasia.

**Figure 6.**
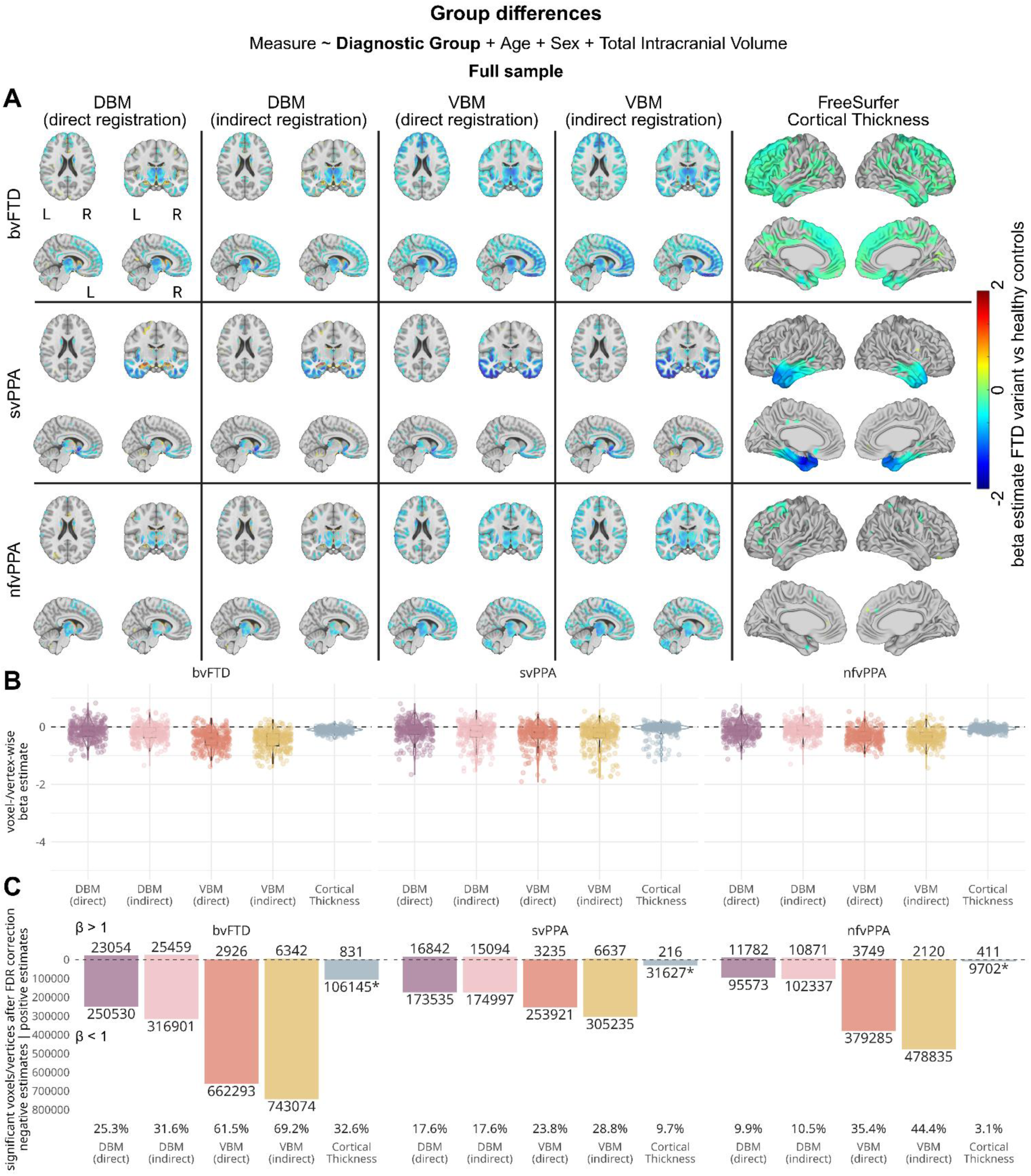
Results of voxel-/vertex-wise linear regression models assessing the sensitivity of each method to detect differences between FTD subtypes and healthy controls in the full sample. A) Brain maps showing beta estimates for the main effect for diagnostic group in the grey matter, comparing voxel-/vertex-wise values for FTD variants versus healthy controls. B) Box- and violin plots summarizing beta estimates for the group effect for each method. Each datapoint represents the estimate for one voxel/vertex. For clearer visualization, 300 datapoints were sampled out of the distribution. C) Barplots showing the number of voxels or vertices with significant group differences, after FDR correction, divided by direction of the effect. Percentages of significant voxels or vertices, regardless of effect direction, are shown below each bar. *Note that Cortical Thickness is calculated for 327,684 vertices while VBM and DBM are extracted for 1,082,282 grey matter voxels. VBM: Voxel-Based Morphometry, DBM: Deformation-Based Morphometry, bvFTD: behavioral-variant Frontotemporal Dementia, svPPA: semantic-variant Primary Progressive Aphasia, nfvPPA: nonfluent-variant Primary Progressive Aphasia.

As DBM and VBM also provide signal for volumetric changes in the white matter, we repeated the voxel-wise analysis in white matter areas for direct and indirect DBM/VBM (Figure 7). Patterns of white matter atrophy matched the locations of grey matter atrophy, whereby bvFTD showed bilateral frontal lobe atrophy, svPPA was focused on the temporal lobes, specifically the left hemisphere, and nfvPPA showed a more diffuse pattern including frontal and temporal lobes (Figure 7A). While indirect VBM generally identified more significant group differences, the performance differences between methods were less pronounced than in grey matter results, and beta estimate ranges were largely similar across methods (Figure 7B/C, for subset results see Supplementary Figure 10, for t-statistics see SUpplementary Figure 11).

**Figure 7.**
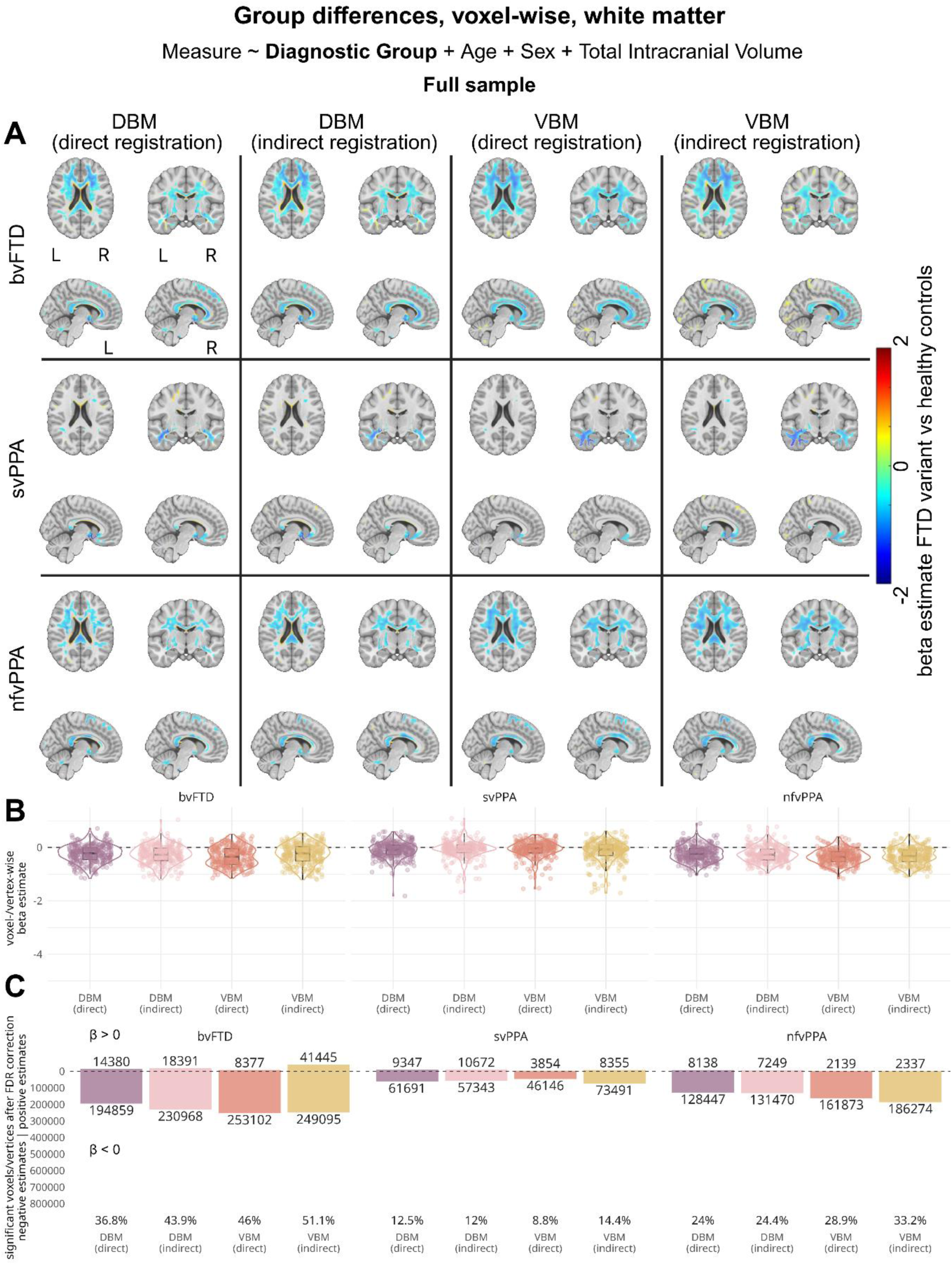
White matter results of voxel--wise linear regression models assessing the sensitivity of each method to detect differences between FTD subtypes and healthy controls in the full sample. A) Brain maps showing beta estimates for the main effect for diagnostic group in the grey matter, comparing voxel-wise values for FTD variants versus healthy controls. B) Box- and violin plots summarizing beta estimates for the group effect for each method. Each datapoint represents the estimate for one voxel. For clearer visualization, 300 datapoints were sampled out of the distribution. C) Barplots showing the number of voxels with significant group differences, after FDR correction, divided by direction of the effect. Percentages of significant voxels, regardless of effect direction, are shown below each bar. VBM: Voxel-Based Morphometry, DBM: Deformation-Based Morphometry, bvFTD: behavioral-variant Frontotemporal Dementia, svPPA: semantic-variant Primary Progressive Aphasia, nfvPPA: nonfluent-variant Primary Progressive Aphasia.

### 3.4. Sensitivity to temporal change in brain atrophy

The sensitivity of each method for detecting longitudinal anatomical changes was assessed by comparing slope differences between FTD variants and healthy controls using linear mixed-effects models. The estimated slopes essentially indicate the increased rates of change for each FTD group that is above and beyond the estimated rate of change in the control group. In the regional analysis of the subset, VBM produced the largest effect sizes (e.g. direct VBM: range(bvFTD) = [-0.14,0.01], range(svPPA) = [-0.47,0.00], range(nfvPPA) = [-0.22,0.02], Figure 8A/B, for t-statistics see Supplementary Figures S12/S13) for most FTD groups, with the exception of svPPA, where CT detected the greatest slope differences in temporal cortex regions (e.g., beta(right entorhinal cortex, svPPA) = - 0.93, p < 0.001). VBM again detected the highest number of significantly different regions after FDR correction (e.g. direct VBM n(bvFTD) = 56, n(svPPA) = 50, n(nfvPPA) = 48, Figure 8C), followed by DBM (e.g. direct DBM n(bvFTD) = 39, n(svPPA) = 49, n(nfvPPA) = 41) and FreeSurfer Volume (n(bvFTD) = 45, n(svPPA) = 47, n(nfvPPA) = 38). DBM exhibited some positive estimates, indicating less atrophy over time in patients versus controls, predominantly in periventricular regions such as the hippocampus (beta(left hippocampus, svPPA = 0.13, p < 0.001). When analyzing the full sample (Figure 9A/B), these patterns persisted. The larger sample size and corresponding increase in statistical power allowed DBM, VBM an FreeSurfer Volume to detect a greater number of significant regions, with the most pronounced gains observed in bvFTD and svPPA (Figure 8C/9C).

**Figure 8.**
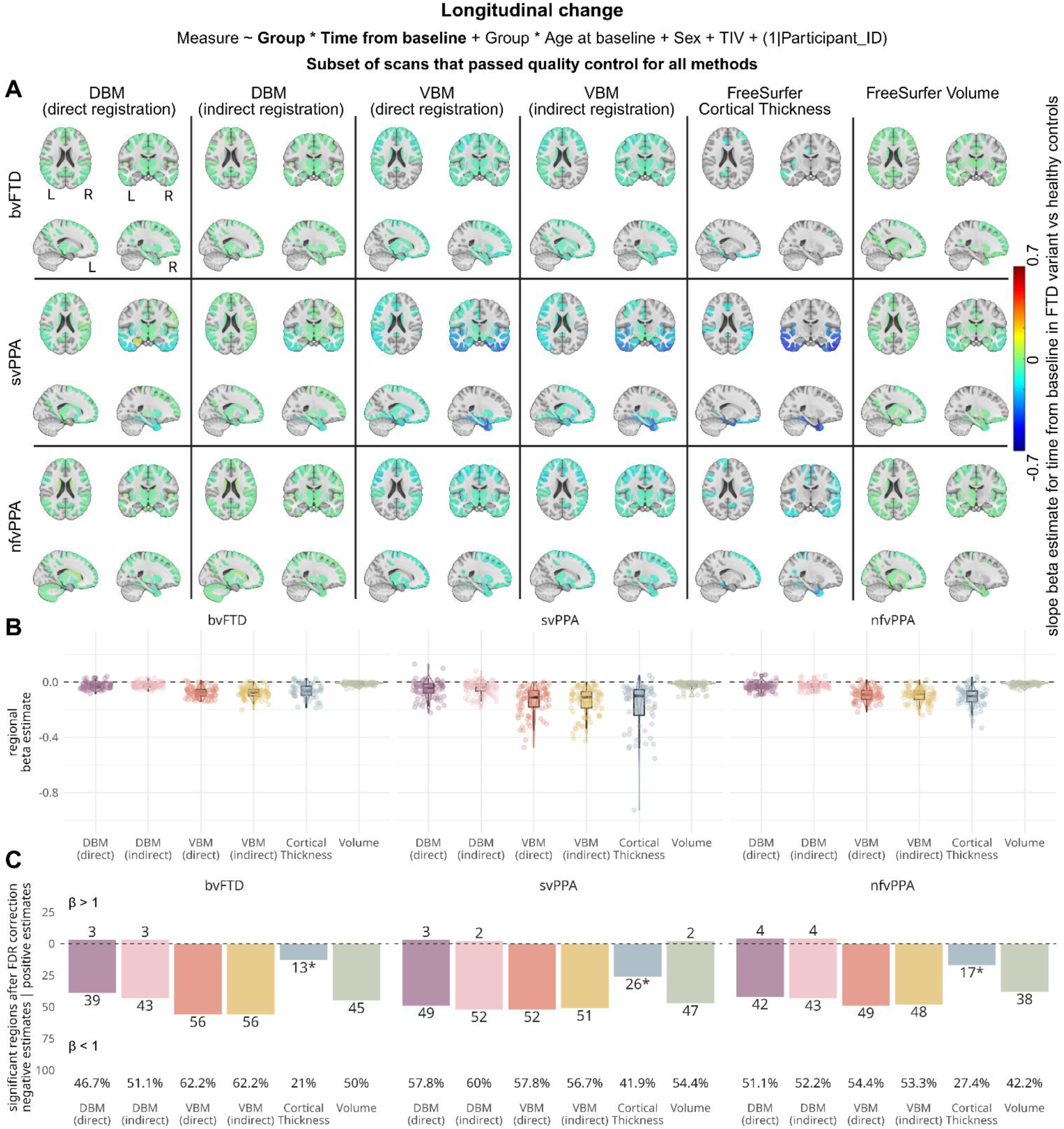
Results of linear mixed-effects models assessing the sensitivity of each method to detect longitudinal changes between FTD subtypes and healthy controls in the subset of MRI scans that were successfully processed by each method. A) Brain maps showing beta estimates for the interaction between diagnostic group and time from baseline, comparing regional slopes for FTD variants versus healthy controls. B) Box- and violin plots summarizing beta estimates for the interaction effect for each method. Each datapoint represents the estimate for one atlas region. C) Barplots showing the number of regions with significant slope differences, after FDR correction, divided by direction of the effect. Percentages of significant regions out of all atlas regions, regardless of effect direction, are shown below each bar. *Note that Cortical Thickness is only calculated for 62 of 90 regions, as this measure does not include subcortical areas. VBM: Voxel-Based Morphometry, DBM: Deformation-Based Morphometry, bvFTD: behavioral-variant Frontotemporal Dementia, svPPA: semantic-variant Primary Progressive Aphasia, nfvPPA: nonfluent-variant Primary Progressive Aphasia.

**Figure 9.**
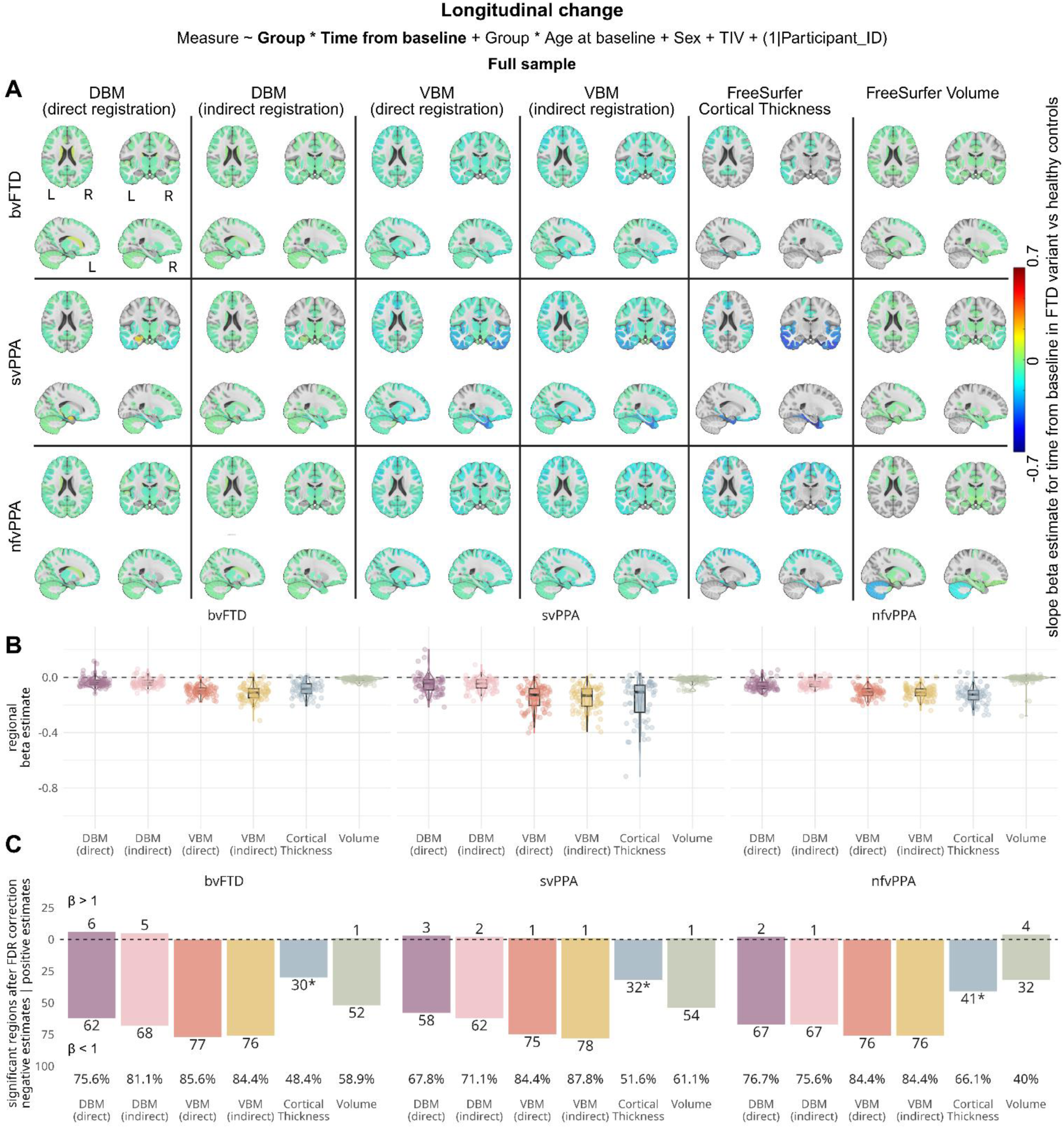
Results of linear mixed-effects models assessing the sensitivity of each method to detect longitudinal changes between FTD subtypes and healthy controls in the full sample. A) Brain maps showing beta estimates for the interaction between diagnostic group and time from baseline, comparing regional slopes for FTD variants versus healthy controls. B) Box- and violin plots summarizing beta estimates for the interaction effect for each method. Each datapoint represents the estimate for one atlas region. C) Barplots showing the number of regions with significant slope differences, after FDR correction, divided by direction of the effect. Percentages of significant regions out of all atlas regions, regardless of effect direction, are shown below each bar. *Note that Cortical Thickness is only calculated for 62 of 90 regions, as this measure does not include subcortical areas. VBM: Voxel-Based Morphometry, DBM: Deformation-Based Morphometry, bvFTD: behavioral-variant Frontotemporal Dementia, svPPA: semantic-variant Primary Progressive Aphasia, nfvPPA: nonfluent-variant Primary Progressive Aphasia.

## 4. Discussion

In this study, we conducted a systematic comparison between commonly applied neuroimaging methods and pipelines for estimating brain morphometry in an FTD cohort. First, pipeline robustness was assessed through rigorous visual quality control at each processing stage using consistent quality control images to identify processing failures. We found that failure rates increased with neurodegenerative and cerebrovascular lesion load in all methods, with notably higher failure rates observed in FreeSurfer. Errors occurred predominantly in areas with advanced atrophy or WMHs. Subsequently, we evaluated the sensitivity of each method for identifying anatomical differences between FTD subtypes and healthy controls, as well as detecting structural changes over time. While all methods revealed broadly consistent patterns of atrophy, VBM produced the highest effect sizes overall. Both DBM and VBM demonstrated superior statistical power for identifying subtle anatomical differences, attributable to lower processing failure rates and consequently larger sample sizes available for group-level analyses.

While PELICAN produced relatively robust outcomes (successful processing for direct DBM: 93.17%, indirect DBM: 97.95%, direct VBM: 91.80%, indirect VBM: 93.17%), FreeSurfer, despite being among the most popular choices, exhibited substantial failure rates (successful processing for Volumes: 77.53%, CT: 61.90%), leading to the exclusion of approximately one-third of scans from downstream analyses. This aligns with prior literature reporting high failure rates even in middle-aged populations (Monereo-Sánchez et al., 2021). Importantly, failure rates were significantly associated with disease severity and atrophy burden, meaning participants with the most pronounced and characteristic disease patterns had to be systematically excluded. These findings indicate that FreeSurfer has limited performance in populations expected to deviate substantially from the healthy templates that the software was trained on, including aging cohorts and dementia patients.

We observed that failure patterns predominantly corresponded to atrophy patterns, whereby higher failure rates occurred in frontal lobe regions for bvFTD and temporal lobe regions for svPPA. Similarly, Raamana et al. (2022) documented errors primarily in the temporal pole, with pial surface underestimation in 35% of cases and 80-100% failure rates in the entorhinal cortex. Vahermaa et al. (2023) likewise identified errors near the middle cerebral artery in 25.2% of scans. Collectively, these results suggest that FreeSurfer should be used cautiously in populations with extensive temporal lobe atrophy, such as FTD and Alzheimer’s disease. Conversely, errors in healthy controls most frequently involved the overestimation of cortical thickness. FreeSurfer’s tendency towards oversegmentation, particularly in larger structures, has been previously documented and attributed to its probabilistic template being derived from young, healthy individuals, making it less representative of elderly or disease populations (Pipitone et al., 2014; Vahermaa et al., 2023; Zandifar et al., 2017). This cumulative evidence suggests that FreeSurfer introduces systematic bias, underestimating measurements in atrophied regions while overestimating them in controls, thereby inflating between-group differences. Hence, meticulous quality control and, where feasible, manual correction are essential when working with FreeSurfer outputs.

For PELICAN, we observed improved robustness when incorporating a population-specific template as an intermediate step during nonlinear registration to standard ICBM space. Although this approach slightly increased errors in healthy controls, the exclusion numbers remained sufficiently low to justify this trade-off, particularly given adequate control sample sizes. Similar findings have been reported in VBM studies, where custom templates and tissue priors enhanced performance (Senjem et al., 2005). Given the prevalence of extensive atrophy in FTD, the use of disease-specific templates is therefore advisable for FTD cohorts. Similarly, tissue segmentation accuracy in PELICAN likely benefits from the inclusion of WMHs as a separate tissue class, accounting for the high cerebrovascular lesion burden characteristic of FTD. In the between-group and longitudinal analyses, we did not detect substantial differences between direct or indirect nonlinear registration. Effect sizes are mostly comparable, and both techniques capture similar regional and voxel-wise differences. However, we did find that including the FTD-template decreased the effect of partial volume leakage slightly, especially in periventricular areas (Figure 6/7).

Based on previous literature reporting higher effect sizes for relative to volumetric measures (Chen et al., 2024; Winkler et al., 2010), we hypothesized that cortical thickness would provide the largest estimates for detecting cross-sectional group and longitudinal slope differences in the subset of participants successfully processed by all methods. Contrary to this hypothesis, our results suggest that for our cohort, VBM is the more sensitive technique, detecting subtle differences in a greater number of regions and yielding larger effect sizes in the subset analysis. This discrepancy potentially reflects differences in how surface-based and volumetric methods relate to biological processes whereby cortical thinning has been primarily associated with neuronal loss, while VBM seems to captures volumetric changes related to surface area reduction (Winkler et al., 2010). Furthermore, spatial smoothing increases VBM’s power for detecting broader regional atrophy (Ashburner & Friston, 2000), as is present in FTD, whereas surface-based methods are more sensitive to focal cortical thinning (Lerch & Evans, 2005). Method selection should therefore be guided by the specific disease pattern (Goto et al., 2022; Scanlon et al., 2011). A notable finding was that CT consistently produced the largest effect sizes in the temporal lobe of svPPA participants. As mentioned above, this region is particularly error-prone in FreeSurfer, and our quality control revealed that FreeSurfer tended to overestimate atrophy. This might suggest that our global quality control was insufficiently sensitive to detect all errors and that future studies should implement thorough region-wise quality control. While manual correction of errors is an option, some evidence suggests that this additional step does not significantly improve group comparison results (Senjem et al., 2005; Vahermaa et al., 2023). Hence, FreeSurfer-based temporal lobe results should be interpreted with caution. VBM also yielded larger effect sizes than DBM in both regional and voxel-wise analyses. This was expected as spatial smoothing increases statistical power by averaging neighboring voxels, which enhances signal-to-noise ratio and reduces variance from minor registration errors, thereby facilitating the detection of true group differences (Whitwell, 2009). Furthermore, VBM appears to benefit from incorporating tissue probability information, which reduces the impact of partial volume effects. Partial volume effects, whereby signal from larger volumes in neighboring voxels (e.g. lateral ventricles) distorts measurements in adjacent regions (e.g. hippocampus or caudate), represent a primary weakness of DBM and were confirmed in this study. This not only resulted in sparse DBM findings in both cortical and subcortical regions but also produced erroneous effects. For instance, positive DBM estimates indicating increased volumes were observed in tissue around the lateral ventricles in the voxelwise group comparison (Figure 6/7) and in the entorhinal cortex in the longitudinal analysis (Figure 8/9). While using a population appropriate template reduced these partial volume effects, they were not eliminated completely. Therefore, mitigating signal leakage by first identifying tissue type (as done in VBM) appears crucial to ensure accurate results. Surprisingly, VBM detected subtle volumetric changes as well as or better than DBM, contrasting with previous reports of DBM’s higher sensitivity to subtle changes, especially in deep grey matter regions, due to its lack of smoothing (Brunaud et al., 2024; Cardenas et al., 2007; Schwarz & Kašpárek, 2011; Song et al., 2022). Considering that the extent of smoothing should be matched to the expected extent of the biological effect (Ashburner & Friston, 2000; Chung et al., 2001; Whitwell, 2009), our relatively small smoothing kernel (4mm) could indicate that modest smoothing may optimize sensitivity for FTD-related atrophy patterns.

In the full-sample comparison, DBM and VBM exhibited even more enhanced performance compared to cortical thickness, attributable to higher participant retention rates following quality control, particularly given that the added participants were mainly those with more extensive levels of atrophy (Table 3, Supplementary Figure S3). Participant loss due to image processing errors significantly reduces the statistical power to detect smaller effects, which particularly affects studies of rare diseases where sample sizes are already limited. These results underscore that the robustness of processing software should be a critical consideration for studies involving populations with advanced brain atrophy or high lesion burden. A key strength of this study is the use of PELICAN, an open-access neuroimaging pipeline that has been developed and extensively validated for use in multi-centre and multi-scanner datasets of aging and neurodegenerative disease populations (Brzezinski-Rittner et al., 2025; Kamal et al., 2025; Metz et al., 2024; Moqadam et al., 2025). PELICAN was explicitly designed to identify cerebrovascular lesions, thereby providing robust morphometric estimates even in cohorts with substantial pathological burden. While FreeSurfer also segments WMHs as a discrete tissue class, it exhibits higher rates of tissue misclassification, leading to less accurate volume estimations. Moreover, FreeSurfer’s surface tessellation algorithm operates without tissue segmentation priors; hence, grey matter/white matter boundary and pial surface reconstructions are more susceptible to errors in the presence of lesions.

Limitations of this study include the modest sample size, particularly for svPPA and nfvPPA, with short follow-up times and few longitudinal observations per participant. Statistical power may therefore have been insufficient to detect subtle between-group differences or meaningful changes over time, potentially explaining the limited significant findings in the temporal change model. Future studies with larger, well-characterized, and diverse cohorts and extended follow-up periods are needed to validate these findings. Furthermore, we focused our analyses on two processing pipelines: our in-house pipeline and the widely used FreeSurfer. Given that FTD patients exhibit high amounts of cerebrovascular pathology (Dadar, Mahmoud, et al., 2022), we required techniques capable of managing WMHs. Alternative software packages, such as ANTs Atropos (Avants et al., 2009), SPM12 (Friston et al., 2011), or FAST from FSL (Zhang et al., 2001), frequently misclassify WMHs as grey matter (Dadar & Collins, 2021), rendering them suboptimal for populations with substantial vascular burden. Although CAT12 (Gaser et al., 2024) implements separate WMH tissue classes, preliminary processing of a subset of participants with these tools revealed substantial segmentation inaccuracies

(Supplementary Figure S14), and we therefore opted against including these methods. A further limitation is that we evaluated only FreeSurfer version 7.4.1, the most recent release available at the time of analysis. Substantial inter-version variability in FreeSurfer outputs, leading to differences in downstream statistical results, has been previously documented (Filip et al., 2022; Gronenschild et al., 2012; Haddad et al., 2023), limiting the generalizability of our findings to other FreeSurfer versions.

Overall, this study underscores the importance of rigorous quality control in neuroimaging research. We demonstrate that the selection of image processing methodology and pipeline profoundly influences effect sizes and statistical power to detect meaningful between-group differences or longitudinal changes. In the FTD cohort examined here, we found that volumetric measures, i.e., DBM and VBM, in combination with the use of a disease-specific template for nonlinear registration yielded sufficiently robust results to maintain adequate statistical power for capturing atrophy patterns after quality control procedures.

## Data Availability

FTLDNI MRI and clinical measures are available through https://ida.loni.usc.edu/login.jsp. PELICAN and FreeSurfer are available at https://github.com/VANDAlab/Preprocessing_Pipeline and https://surfer.nmr.mgh.harvard.edu/, respectively. Codes used for analysis are available on GitHub (https://github.com/ameliemetz/FTD_methods_comparison).

## Data and Code Availability

FTLDNI MRI and clinical measures are available through https://ida.loni.usc.edu/login.jsp. PELICAN and FreeSurfer are available at https://github.com/VANDAlab/Preprocessing_Pipeline and https://surfer.nmr.mgh.harvard.edu/, respectively. Codes used for analysis are available on GitHub (https://github.com/ameliemetz/FTD_methods_comparison). Regional and voxel-/vertex-wise results are available for download under https://zenodo.org/records/17516941.

## Author Contributions

Amelie Metz: Conceptualization, Visualization, Writing - Original Draft, Formal analysis, Software

Roqaie Moqadam: Visualization, Software

Louis Collins: Conceptualization, Writing - Review & Editing

Yashar Zeighami: Conceptualization, Writing - Review & Editing Sylvia

Villeneuve: Conceptualization, Writing - Review & Editing

Mahsa Dadar: Conceptualization, Supervision, Writing - Review & Editing, Software

## Acknowledgements

The authors acknowledge Digital Research Alliance of Canada (https://www.alliancecan.ca/en) for the usage of the computing resources in the current work. Data collection and sharing for this project was also funded by the Frontotemporal Lobar Degeneration Neuroimaging Initiative (National Institutes of Health Grant R01 AG032306). The study is coordinated through the University of California, San Francisco, Memory and Aging Center. FTLDNI data are disseminated by the Laboratory for Neuro Imaging at the University of Southern California.

## References

Akhmadullina, D. R., Konovalov, R. N., Shpilyukova, Y. A., Nevzorova, K. V., Fedotova, E. Yu., & Illarioshkin, S. N. (2024). Neuroanatomical correlates of language impairment in non-fluent variant of primary progressive aphasia. Frontiers in Human Neuroscience, 18, 1486809. 10.3389/fnhum.2024.1486809

Ashburner, J., & Friston, K. (n.d.). Morphometry. In Statistical Parametric Mapping: The Analysis of Functional Brain Images (2nd ed.).

Ashburner, J., & Friston, K. J. (2000). Voxel-Based Morphometry—The Methods. NeuroImage, 11(6), 805–821. 10.1006/nimg.2000.0582

Ashburner, J., Hutton, C., Frackowiak, R., Johnsrude, I., Price, C., & Friston, K. (1998). Identifying global anatomical differences: Deformation-based morphometry. Human Brain Mapping, 6(5–6), 348–357. 10.1002/(SICI)1097-0193(1998)6:5/6%253C348::AID-HBM4%253E3.0.CO;2-P

Avants, B., Tustison, N., Gee, J., & Song, G. (2009). *ANTS: Advanced Open-Source Normalization Tools for Neuroanatomy* [Computer software]. Penn Image Computing and Science Laboratory.

Bang, J., Spina, S., & Miller, B. L. (2015). Frontotemporal dementia. The Lancet, 386(10004), 1672–1682. 10.1016/S0140-6736(15)00461-4

Benjamini, Y., & Hochberg, Y. (1995). Controlling The False Discovery Rate—A Practical And Powerful Approach To Multiple Testing. Journal of the Royal Statistical Society Series B (Methodological), 57, 289–300. DOI:10.2307/2346101

Bisenius, S., Neumann, J., & Schroeter, M. L. (2016). Validating new diagnostic imaging criteria for primary progressive aphasia via anatomical likelihood estimation meta-analyses. European Journal of Neurology, 23(4), 704–712. 10.1111/ene.12902

Brunaud, C., Valable, S., Ropars, G., Dwiri, F.-A., Naveau, M., Toutain, J., Bernaudin, M., Freret, T., Léger, M., Touzani, O., & Pérès, E. A. (2024). Deformation-based morphometry: A sensitive imaging approach to detect radiation-induced brain injury? Cancer Imaging, 24(1), 95. 10.1186/s40644-024-00736-1

Brzezinski-Rittner, A., Moqadam, R., Zeighami, Y., & Dadar, M. (2025). Brain Size: To Adjust or Not Adjust? It’s Not a Matter of If, but How. 10.1101/2025.09.21.25336298

Cardenas, V. A., Boxer, A. L., Chao, L. L., Gorno-Tempini, M. L., Miller, B. L., Weiner, M. W., & Studholme, C. (2007). Deformation-Based Morphometry Reveals Brain Atrophy in Frontotemporal Dementia. Archives of Neurology, 64(6), 873. 10.1001/archneur.64.6.873

Chan, D., Fox, N. C., Scahill, R. I., Crum, W. R., Whitwell, J. L., Leschziner, G., Rossor, A. M., Stevens, J. M., Cipolotti, L., & Rossor, M. N. (2001). Patterns of temporal lobe atrophy in semantic dementia and Alzheimer’s disease. Annals of Neurology, 49(4), 433–442.

Chen, Z., Xie, Q., Wang, J., Wang, Y., Zhang, H., Li, C., Wang, Y., Cong, L., Tang, S., Hou, T., Song, L., Du, Y., & Qiu, C. (2024). Mapping grey matter and cortical thickness alterations associated with subjective cognitive decline and mild cognitive impairment among rural-dwelling older adults in China: A population-based study. NeuroImage. Clinical, 44, 103691. 10.1016/j.nicl.2024.103691

Chu, M., Jiang, D., Li, D., Yan, S., Liu, L., Nan, H., Wang, Y., Wang, Y., Yue, A., Ren, L., Chen, K., Rosa-Neto, P., Lu, J., & Wu, L. (2024). Atrophy network mapping of clinical subtypes and main symptoms in frontotemporal dementia. *Brain*, awae067. 10.1093/brain/awae067

Chung, M. K., Worsley, K. J., Paus, T., Cherif, C., Collins, D. L., Giedd, J. N., Rapoport, J. L., & Evans, A. C. (2001). A Unified Statistical Approach to Deformation-Based Morphometry. NeuroImage, 14(3), 595–606. 10.1006/nimg.2001.0862

Coupe, P., Yger, P., Prima, S., Hellier, P., Kervrann, C., & Barillot, C. (2008). An Optimized Blockwise Nonlocal Means Denoising Filter for 3-D Magnetic Resonance Images. IEEE Transactions on Medical Imaging, 27(4), 425–441. 10.1109/TMI.2007.906087

Dadar, M., Camicioli, R., & Duchesne, S. (2022). Multi sequence average templates for aging and neurodegenerative disease populations. Scientific Data, 9(1), 238. 10.1038/s41597-022-01341-2

Dadar, M., & Collins, D. L. (2021). BISON: Brain tissue segmentation pipeline using T1 -weighted magnetic resonance images and a random forest classifier. Magnetic Resonance in Medicine, 85(4), 1881–1894. 10.1002/mrm.28547

Dadar, M., Fonov, V. S., & Collins, D. L. (2018). A comparison of publicly available linear MRI stereotaxic registration techniques. NeuroImage, 174, 191–200. 10.1016/j.neuroimage.2018.03.025

Dadar, M., Mahmoud, S., Zhernovaia, M., Camicioli, R., Maranzano, J., & Duchesne, S. (2022). White matter hyperintensity distribution differences in aging and neurodegenerative disease cohorts. NeuroImage: Clinical, 36, 103204. 10.1016/j.nicl.2022.103204

Dadar, M., Manera, A. L., Fonov, V. S., Ducharme, S., & Collins, D. L. (2021). MNI-FTD templates, unbiased average templates of frontotemporal dementia variants. Scientific Data, 8(1), 222. 10.1038/s41597-021-01007-5

Dadar, M., Maranzano, J., Ducharme, S., Carmichael, O. T., Decarli, C., Collins, D. L., & Alzheimer’s Disease Neuroimaging Initiative. (2018). Validation of T 1w-based segmentations of white matter hyperintensity volumes in large-scale datasets of aging. Human Brain Mapping, 39(3), 1093–1107. 10.1002/hbm.23894

Dadar, M., Moqadam, R., Metz, A., Chadwick, K., Brzezinski-Rittner, A., & Zeighami, Y. (2025). PELICAN: A Longitudinal Image Processing Pipeline for Analyzing Structural Magnetic Resonance Images in Aging and Neurodegenerative Disease Populations. 10.1101/2025.09.20.677546

Dadar, M., Potvin, O., Camicioli, R., Duchesne, S., & for the Alzheimer’s Disease Neuroimaging Initiative. (2021). Beware of white matter hyperintensities causing systematic errors in FREESURFER gray matter segmentations! Human Brain Mapping, 42(9), 2734–2745. 10.1002/hbm.25398

Dale, A. M., Fischl, B., & Sereno, M. I. (1999). Cortical Surface-Based Analysis. NeuroImage, 9(2), 179–194. 10.1006/nimg.1998.0395

Davies, R. R., Halliday, G. M., Xuereb, J. H., Kril, J. J., & Hodges, J. R. (2009). The neural basis of semantic memory: Evidence from semantic dementia. Neurobiology of Aging, 30(12), 2043–2052. 10.1016/j.neurobiolaging.2008.02.005

Desikan, R. S., Ségonne, F., Fischl, B., Quinn, B. T., Dickerson, B. C., Blacker, D., Buckner, R. L., Dale, A. M., Maguire, R. P., Hyman, B. T., Albert, M. S., & Killiany, R. J. (2006). An automated labeling system for subdividing the human cerebral cortex on MRI scans into gyral based regions of interest. NeuroImage, 31(3), 968–980. 10.1016/j.neuroimage.2006.01.021

Eskildsen, S. F., Coupé, P., Fonov, V., Manjón, J. V., Leung, K. K., Guizard, N., Wassef, S. N., Østergaard, L. R., & Collins, D. L. (2012). BEaST: Brain extraction based on nonlocal segmentation technique. NeuroImage, 59(3), 2362–2373. 10.1016/j.neuroimage.2011.09.012

Fereshtehnejad, S., Moqadam, R., Azizi, H., Postuma, R. B., Dadar, M., Lang, A. E., Marras, C., & Zeighami, Y. (2025). Distinct Longitudinal Clinical-Neuroanatomical Trajectories in Parkinson’s Disease Clinical Subtypes: Insight toward Precision Medicine. Movement Disorders, 40(8), 1572–1583. 10.1002/mds.30229

Filip, P., Bednarik, P., Eberly, L. E., Moheet, A., Svatkova, A., Grohn, H., Kumar, A. F., Seaquist, E. R., & Mangia, S. (2022). Different FreeSurfer versions might generate different statistical outcomes in case–control comparison studies. Neuroradiology, 64(4), 765–773. 10.1007/s00234-021-02862-0

Fischl, B. (2012). FreeSurfer. NeuroImage, 62(2), 774–781. 10.1016/j.neuroimage.2012.01.021

Fischl, B., Salat, D. H., Busa, E., Albert, M., Dieterich, M., Haselgrove, C., Van Der Kouwe, A., Killiany, R., Kennedy, D., Klaveness, S., Montillo, A., Makris, N., Rosen, B., & Dale, A. M. (2002). Whole Brain Segmentation. Neuron, 33(3), 341–355. 10.1016/S0896-6273(02)00569-X

Fischl, B., Sereno, M. I., & Dale, A. M. (1999). Cortical Surface-Based Analysis. NeuroImage, 9(2), 195–207. 10.1006/nimg.1998.0396

Fonov, V., Evans, A. C., Botteron, K., Almli, C. R., McKinstry, R. C., & Collins, D. L. (2011). Unbiased average age-appropriate atlases for pediatric studies. NeuroImage, 54(1), 313–327. 10.1016/j.neuroimage.2010.07.033

Friston, K. J., Ashburner, J. T., Kiebel, S., Nichols, T. E., & Penny, W. D. (2011). Statistical Parametric Mapping: The Analysis of Functional Brain Images. Elsevier Science.

Galton, C. J., Patterson, K., Graham, K., Lambon-Ralph, M. A., Williams, G., Antoun, N., Sahakian, B. J., & Hodges, J. R. (2001). Differing patterns of temporal atrophy in Alzheimer’s disease and semantic dementia. Neurology, 57(2), 216–225. 10.1212/WNL.57.2.216

Gaser, C., Dahnke, R., Thompson, P. M., Kurth, F., Luders, E., & the Alzheimer’s Disease Neuroimaging Initiative. (2024). CAT: A computational anatomy toolbox for the analysis of structural MRI data. GigaScience, 13, giae049. 10.1093/gigascience/giae049

Genovese, C. R., Lazar, N. A., & Nichols, T. (2002). Thresholding of Statistical Maps in Functional Neuroimaging Using the False Discovery Rate. NeuroImage, 15(4), 870–878. 10.1006/nimg.2001.1037

Gorno-Tempini, M. L., Hillis, A. E., Weintraub, S., Kertesz, A., Mendez, M., Cappa, S. F., Ogar, J. M., Rohrer, J. D., Black, S., Boeve, B. F., Manes, F., Dronkers, N. F., Vandenberghe, R., Rascovsky, K., Patterson, K., Miller, B. L., Knopman, D. S., Hodges, J. R., Mesulam, M. M., & Grossman, M. (2011). Classification of primary progressive aphasia and its variants. Neurology, 76(11), 1006–1014. 10.1212/WNL.0b013e31821103e6

Goto, M., Abe, O., Hagiwara, A., Fujita, S., Kamagata, K., Hori, M., Aoki, S., Osada, T., Konishi, S., Masutani, Y., Sakamoto, H., Sakano, Y., Kyogoku, S., & Daida, H. (2022). Advantages of Using Both Voxel- and Surface-based Morphometry in Cortical Morphology Analysis: A Review of Various Applications. Magnetic Resonance in Medical Sciences, 21(1), 41–57. 10.2463/mrms.rev.2021-0096

Gronenschild, E. H. B. M., Habets, P., Jacobs, H. I. L., Mengelers, R., Rozendaal, N., Van Os, J., & Marcelis, M. (2012). The Effects of FreeSurfer Version, Workstation Type, and Macintosh Operating System Version on Anatomical Volume and Cortical Thickness Measurements. PLoS ONE, 7(6), e38234. 10.1371/journal.pone.0038234

Haddad, E., Pizzagalli, F., Zhu, A. H., Bhatt, R. R., Islam, T., Ba Gari, I., Dixon, D., Thomopoulos, S. I., Thompson, P. M., & Jahanshad, N. (2023). Multisite test-retest reliability and compatibility of brain metrics derived from FreeSurfer versions 7.1, 6.0, and 5.3. Human Brain Mapping, 44(4), 1515–1532. 10.1002/hbm.26147

Kamal, F., Moqadam, R., Morrison, C., & Dadar, M. (2025). Racial and ethnic differences in white matter hypointensities: The role of vascular risk factors. Alzheimer’s & Dementia, 21(3), e70105. 10.1002/alz.70105

King, D. J., Novak, J., Shephard, A. J., Beare, R., Anderson, V. A., & Wood, A. G. (2020). Lesion Induced Error on Automated Measures of Brain Volume: Data From a Pediatric Traumatic Brain Injury Cohort. Frontiers in Neuroscience, 14, 491478. 10.3389/fnins.2020.491478

Klasson, N., Olsson, E., Eckerström, C., Malmgren, H., & Wallin, A. (2018). Estimated intracranial volume from FreeSurfer is biased by total brain volume. European Radiology Experimental, 2(1), 24. 10.1186/s41747-018-0055-4

Kurth, F., Luders, E., & Gaser, C. (2015). Voxel-Based Morphometry. In Brain Mapping (pp. 345–349). Elsevier. 10.1016/B978-0-12-397025-1.00304-3

Lajoie, I., Canadian ALS Neuroimaging Consortium (CALSNIC), Kalra, S., & Dadar, M. (2025). Regional Cerebral Atrophy Contributes to Personalized Survival Prediction in Amyotrophic Lateral Sclerosis: A Multicentre, Machine Learning, Deformation-Based Morphometry Study. Annals of Neurology, 97(6), 1144–1157. 10.1002/ana.27196

Lerch, J. P. (2015). Cortical Thickness Mapping. In Brain Mapping (pp. 351–355). Elsevier. 10.1016/B978-0-12-397025-1.00305-5

Lerch, J. P., & Evans, A. C. (2005). Cortical thickness analysis examined through power analysis and a population simulation. NeuroImage, 24(1), 163–173. 10.1016/j.neuroimage.2004.07.045

Mackenzie, I. R. A., Neumann, M., Bigio, E. H., Cairns, N. J., Alafuzoff, I., Kril, J., Kovacs, G. G., Ghetti, B., Halliday, G., Holm, I. E., Ince, P. G., Kamphorst, W., Revesz, T., Rozemuller, A. J. M., Kumar-Singh, S., Akiyama, H., Baborie, A., Spina, S., Dickson, D. W., … Mann, D. M. A. (2009). Nomenclature for neuropathologic subtypes of frontotemporal lobar degeneration: Consensus recommendations. Acta Neuropathologica, 117(1), 15–18. 10.1007/s00401-008-0460-5

Mackenzie, I. R. A., Neumann, M., Bigio, E. H., Cairns, N. J., Alafuzoff, I., Kril, J., Kovacs, G. G., Ghetti, B., Halliday, G., Holm, I. E., Ince, P. G., Kamphorst, W., Revesz, T., Rozemuller, A. J. M., Kumar-Singh, S., Akiyama, H., Baborie, A., Spina, S., Dickson, D. W., … Mann, D. M. A. (2010). Nomenclature and nosology for neuropathologic subtypes of frontotemporal lobar degeneration: An update. Acta Neuropathologica, 119(1), 1–4. 10.1007/s00401-009-0612-2

Manera, A. L., Dadar, M., Collins, D. L., & Ducharme, S. (2019). Deformation based morphometry study of longitudinal MRI changes in behavioral variant frontotemporal dementia. NeuroImage: Clinical, 24, 102079. 10.1016/j.nicl.2019.102079

Manera, A. L., Dadar, M., Collins, D. L., & Ducharme, S. (2022). Ventricular features as reliable differentiators between bvFTD and other dementias. NeuroImage: Clinical, 33, 102947. 10.1016/j.nicl.2022.102947

Manera, A. L., Dadar, M., Fonov, V., & Collins, D. L. (2020). CerebrA, registration and manual label correction of Mindboggle-101 atlas for MNI-ICBM152 template. Scientific Data, 7(1), 237. 10.1038/s41597-020-0557-9

Mechelli, A., Price, C., Friston, K., & Ashburner, J. (2005). Voxel-Based Morphometry of the Human Brain: Methods and Applications. Current Medical Imaging Reviews, 1(2), 105–113. 10.2174/1573405054038726

Meeter, L. H., Kaat, L. D., Rohrer, J. D., & Van Swieten, J. C. (2017). Imaging and fluid biomarkers in frontotemporal dementia. Nature Reviews Neurology, 13(7), 406–419. 10.1038/nrneurol.2017.75

Mesulam, M., Wieneke, C., Rogalski, E., Cobia, D., Thompson, C., & Weintraub, S. (2009). Quantitative Template for Subtyping Primary Progressive Aphasia. Archives of Neurology, 66(12). 10.1001/archneurol.2009.288

Metz, A., Zeighami, Y., Ducharme, S., Villeneuve, S., & Dadar, M. (2024). Frontotemporal dementia subtyping using machine learning, multivariate statistics and neuroimaging. Brain Communications, 7(1), fcaf065. 10.1093/braincomms/fcaf065

Monereo-Sánchez, J., De Jong, J. J. A., Drenthen, G. S., Beran, M., Backes, W. H., Stehouwer, C. D. A., Schram, M. T., Linden, D. E. J., & Jansen, J. F. A. (2021). Quality control strategies for brain MRI segmentation and parcellation: Practical approaches and recommendations - insights from the Maastricht study. NeuroImage, 237, 118174. 10.1016/j.neuroimage.2021.118174

Moqadam, R., Azizi, H., Brzezinski-Rittner, A., Ronat, L. A., Raeesi, S., Hanganu, A., Zeighami, Y., & Dadar, M. (2025). Apathy progression is associated with brain atrophy and white matter damage in Parkinson’s disease. *Brain Communications*, fcaf355. 10.1093/braincomms/fcaf355

Moqadam, R., Dadar, M., & Zeighami, Y. (2024). Investigating the impact of motion in the scanner on brain age predictions. Imaging Neuroscience, 2, imag–2–00079. 10.1162/imag_a_00079

Neelin, P., MacDonald, D., Collins, D. L., & Evans, A. C. (1998). The MINC file format: From bytes to brains. NeuroImage, 7(4), S786. 10.1016/S1053-8119(18)31619-7

Olney, N. T., Spina, S., & Miller, B. L. (2017). Frontotemporal Dementia. Neurologic Clinics, 35(2), 339–374. 10.1016/j.ncl.2017.01.008

Pan, P. L., Song, W., Yang, J., Huang, R., Chen, K., Gong, Q. Y., Zhong, J. G., Shi, H. C., & Shang, H. F. (2012). Gray Matter Atrophy in Behavioral Variant Frontotemporal Dementia: A Meta-Analysis of Voxel-Based Morphometry Studies. Dementia and Geriatric Cognitive Disorders, 33(2–3), 141–148. 10.1159/000338176

Pipitone, J., Park, M. T. M., Winterburn, J., Lett, T. A., Lerch, J. P., Pruessner, J. C., Lepage, M., Voineskos, A. N., & Chakravarty, M. M. (2014). Multi-atlas segmentation of the whole hippocampus and subfields using multiple automatically generated templates. NeuroImage, 101, 494–512. 10.1016/j.neuroimage.2014.04.054

Planche, V., Mansencal, B., Manjon, J. V., Tourdias, T., Catheline, G., Coupé, P., & For the Frontotemporal Lobar Degeneration Neuroimaging Initiative and the National Alzheimer’s Coordinating Center cohort. (2023). Anatomical MRI staging of frontotemporal dementia variants. Alzheimer’s & Dementia, 19(8), 3283–3294. 10.1002/alz.12975

Qiu, T., Liu, Z., Rheault, F., Legarreta, J. H., Valcourt Caron, A., St-Onge, F., Strikwerda-Brown, C., Metz, A., Dadar, M., Soucy, J., Pichet Binette, A., Spreng, R. N., Descoteaux, M., Villeneuve, S., & for the PREVENT-AD Research Group. (2024). Structural white matter properties and cognitive resilience to tau pathology. Alzheimer’s & Dementia, 20(5), 3364–3377. 10.1002/alz.13776

Raamana, P. R., Theyers, A., Selliah, T., Bhati, P., Arnott, S. R., Hassel, S., Nanayakkara, N. D., M. Scott, C. J., Harris, J., Zamyadi, M., Lam, R. W., Milev, R., Müller, D. J., Rotzinger, S., Frey, B. N., Kennedy, S. H., Black, S. E., Lang, A., Masellis, M., … C. Strother, S. (2022). Visual QC Protocol for FreeSurfer Cortical Parcellations from Anatomical MRI. Aperture Neuro, 76. 10.52294/1cdce19c-e6db-4684-97cb-ae709da06a3f

Rademakers, R., Neumann, M., & Mackenzie, I. R. (2012). Advances in understanding the molecular basis of frontotemporal dementia. Nature Reviews Neurology, 8(8), 423–434. 10.1038/nrneurol.2012.117

Rascovsky, K., Hodges, J. R., Knopman, D., Mendez, M. F., Kramer, J. H., Neuhaus, J., Van Swieten, J. C., Seelaar, H., Dopper, E. G. P., Onyike, C. U., Hillis, A. E., Josephs, K. A., Boeve, B. F., Kertesz, A., Seeley, W. W., Rankin, K. P., Johnson, J. K., Gorno-Tempini, M.-L., Rosen, H., … Miller, B. L. (2011). Sensitivity of revised diagnostic criteria for the behavioural variant of frontotemporal dementia. Brain, 134(9), 2456–2477. 10.1093/brain/awr179

Reuter, M., Schmansky, N. J., Rosas, H. D., & Fischl, B. (2012). Within-subject template estimation for unbiased longitudinal image analysis. NeuroImage, 61(4), 1402–1418. 10.1016/j.neuroimage.2012.02.084

Righart, R., Schmidt, P., Dahnke, R., Biberacher, V., Beer, A., Buck, D., Hemmer, B., Kirschke, J. S., Zimmer, C., Gaser, C., & Mühlau, M. (2017). Volume versus surface-based cortical thickness measurements: A comparative study with healthy controls and multiple sclerosis patients. PLOS ONE, 12(7), e0179590. 10.1371/journal.pone.0179590

Rogalski, E., Cobia, D., Martersteck, A., Rademaker, A., Wieneke, C., Weintraub, S., & Mesulam, M.-M. (2014). Asymmetry of cortical decline in subtypes of primary progressive aphasia. Neurology, 83(13), 1184–1191. 10.1212/WNL.0000000000000824

Rosen, H. J., Allison, S. C., Schauer, G. F., Gorno-Tempini, M. L., Weiner, M. W., & Miller, B. L. (2005). Neuroanatomical correlates of behavioural disorders in dementia. Brain, 128(11), 2612–2625. 10.1093/brain/awh628

Scanlon, C., Mueller, S. G., Tosun, D., Cheong, I., Garcia, P., Barakos, J., Weiner, M. W., & Laxer, K. D. (2011). Impact of Methodologic Choice for Automatic Detection of Different Aspects of Brain Atrophy by Using Temporal Lobe Epilepsy as a Model. American Journal of Neuroradiology, 32(9), 1669–1676. 10.3174/ajnr.A2578

Schroeter, M. L., Laird, A. R., Chwiesko, C., Deuschl, C., Schneider, E., Bzdok, D., Eickhoff, S. B., & Neumann, J. (2014). Conceptualizing neuropsychiatric diseases with multimodal data-driven meta-analyses – The case of behavioral variant frontotemporal dementia. Cortex, 57, 22–37. 10.1016/j.cortex.2014.02.022

Schwarz, D., & Kašpárek, T. (2011). Comparison of Two Methods for Automatic Brain Morphometry Analysis. RadioEngineering, 20(4), 996–1001.

Seeley, W. W., Crawford, R., Rascovsky, K., Kramer, J. H., Weiner, M., Miller, B. L., & Gorno-Tempini, M. L. (2008). Frontal Paralimbic Network Atrophy in Very Mild Behavioral Variant Frontotemporal Dementia. Archives of Neurology, 65(2). 10.1001/archneurol.2007.38

Senjem, M. L., Gunter, J. L., Shiung, M. M., Petersen, R. C., & Jack, C. R. (2005). Comparison of different methodological implementations of voxel-based morphometry in neurodegenerative disease. NeuroImage, 26(2), 600–608. 10.1016/j.neuroimage.2005.02.005

Shafiei, G., Bazinet, V., Dadar, M., Manera, A. L., Collins, D. L., Dagher, A., Borroni, B., Sanchez-Valle, R., Moreno, F., Laforce, R., Graff, C., Synofzik, M., Galimberti, D., Rowe, J. B., Masellis, M., Tartaglia, M. C., Finger, E., Vandenberghe, R., De Mendonça, A., … Polyakova, M. (2023). Network structure and transcriptomic vulnerability shape atrophy in frontotemporal dementia. Brain, 146(1), 321–336. 10.1093/brain/awac069

Sled, J. G., Zijdenbos, A. P., & Evans, A. C. (1998). A nonparametric method for automatic correction of intensity nonuniformity in MRI data. IEEE Transactions on Medical Imaging, 17(1), 87–97. 10.1109/42.668698

Song, Z., Krishnan, A., Gaetano, L., Tustison, N. J., Clayton, D., De Crespigny, A., Bengtsson, T., Jia, X., & Carano, R. A. D. (2022). Deformation-based morphometry identifies deep brain structures protected by ocrelizumab. NeuroImage: Clinical, 34, 102959. 10.1016/j.nicl.2022.102959

Vahermaa, V., Aydogan, D. B., Raij, T., Armio, R.-L., Laurikainen, H., Saramäki, J., & Suvisaari, J. (2023). FreeSurfer 7 quality control: Key problem areas and importance of manual corrections. NeuroImage, 279, 120306. 10.1016/j.neuroimage.2023.120306

Vincent, R. D., Neelin, P., Khalili-Mahani, N., Janke, A. L., Fonov, V. S., Robbins, S. M., Baghdadi, L., Lerch, J., Sled, J. G., Adalat, R., MacDonald, D., Zijdenbos, A. P., Collins, D. L., & Evans, A. C. (2016). MINC 2.0: A Flexible Format for Multi-Modal Images. Frontiers in Neuroinformatics, 10. 10.3389/fninf.2016.00035

Whitwell, J. L. (2009). Voxel-Based Morphometry: An Automated Technique for Assessing Structural Changes in the Brain. The Journal of Neuroscience, 29(31), 9661–9664. 10.1523/JNEUROSCI.2160-09.2009

Winkler, A. M., Kochunov, P., Blangero, J., Almasy, L., Zilles, K., Fox, P. T., Duggirala, R., & Glahn, D. C. (2010). Cortical thickness or grey matter volume? The importance of selecting the phenotype for imaging genetics studies. NeuroImage, 53(3), 1135–1146. 10.1016/j.neuroimage.2009.12.028

Wisse, L. E. M., Ungrady, M. B., Ittyerah, R., Lim, S. A., Yushkevich, P. A., Wolk, D. A., Irwin, D. J., Das, S. R., & Grossman, M. (2021). Cross-sectional and longitudinal medial temporal lobe subregional atrophy patterns in semantic variant primary progressive aphasia. Neurobiology of Aging, 98, 231–241. 10.1016/j.neurobiolaging.2020.11.012

Zandifar, A., Fonov, V., Coupé, P., Pruessner, J., & Collins, D. L. (2017). A comparison of accurate automatic hippocampal segmentation methods. NeuroImage, 155, 383–393. 10.1016/j.neuroimage.2017.04.018

Zhang, Y., Brady, M., & Smith, S. (2001). Segmentation of brain MR images through a hidden Markov random field model and the expectation-maximization algorithm. IEEE Transactions on Medical Imaging, 20(1), 45–57. 10.1109/42.906424

