## Supplementary Material for "Quantifying brain atrophy in Frontotemporal Dementia: a head-to-head comparison of neuroimaging techniques"

### Supplementary Materials

Metz et al.

Quantifying brain atrophy in Frontotemporal Dementia:

a head-to-head comparison of neuroimaging techniques

#### Supplementary Table S1: MRI acquisition parameters and Number of participants recruited, scanned, and tested at each of the FTLDNI research sites

| | **controls** | **bvFTD** | **svPPA** | **nfvPPA** |
|---|---|---|---|---|
| *Mayo* | 9 | 16 | 1 | 0 |
| *MGH* | 0 | 5 | 4 | 3 |
| *UCSF* | 127 | 56 | 34 | 37 |

bvFTD: behavioral-variant Frontotemporal Dementia, svPPA: semantic-variant Primary Progressive Aphasia, nfvPPA: non-fluent variant Primary Progressive Aphasia.

3.0 T MRIs were acquired at three sites [T1-weighted magnetization-prepared rapid gradient-echo imaging (MPRAGE)] using Siemens Trio Total imaging matrix (Tim) scanners, with the following parameters:

- UCSF: repetition time (TR) = 2.3 ms, echo time = 2.98 ms, inversion time = 900 ms, flip angle 9°, matrix 256 × 240, slice thickness 1 mm, voxel size 1 mm3
- Mayo: repetition time (TR) = 2.3 ms, echo time = 3.04 ms, inversion time = 900 ms, flip angle 8°, matrix 256 × 240, slice thickness 1.2 mm, voxel size 1 mm3
- MGH: repetition time (TR) = 2.3 ms, echo time = 2.96 ms, inversion time = 900 ms, flip angle 9°, matrix 256 × 240, slice thickness 1 mm, voxel size 1 mm3

The FTLDNI uses the infrastructure established by the Alzheimer's Disease Neuroimaging Initiative (ADNI). All participating imaging centers share a common platform.

#### Supplementary Figure S2: Flow chart indicating number of participants included

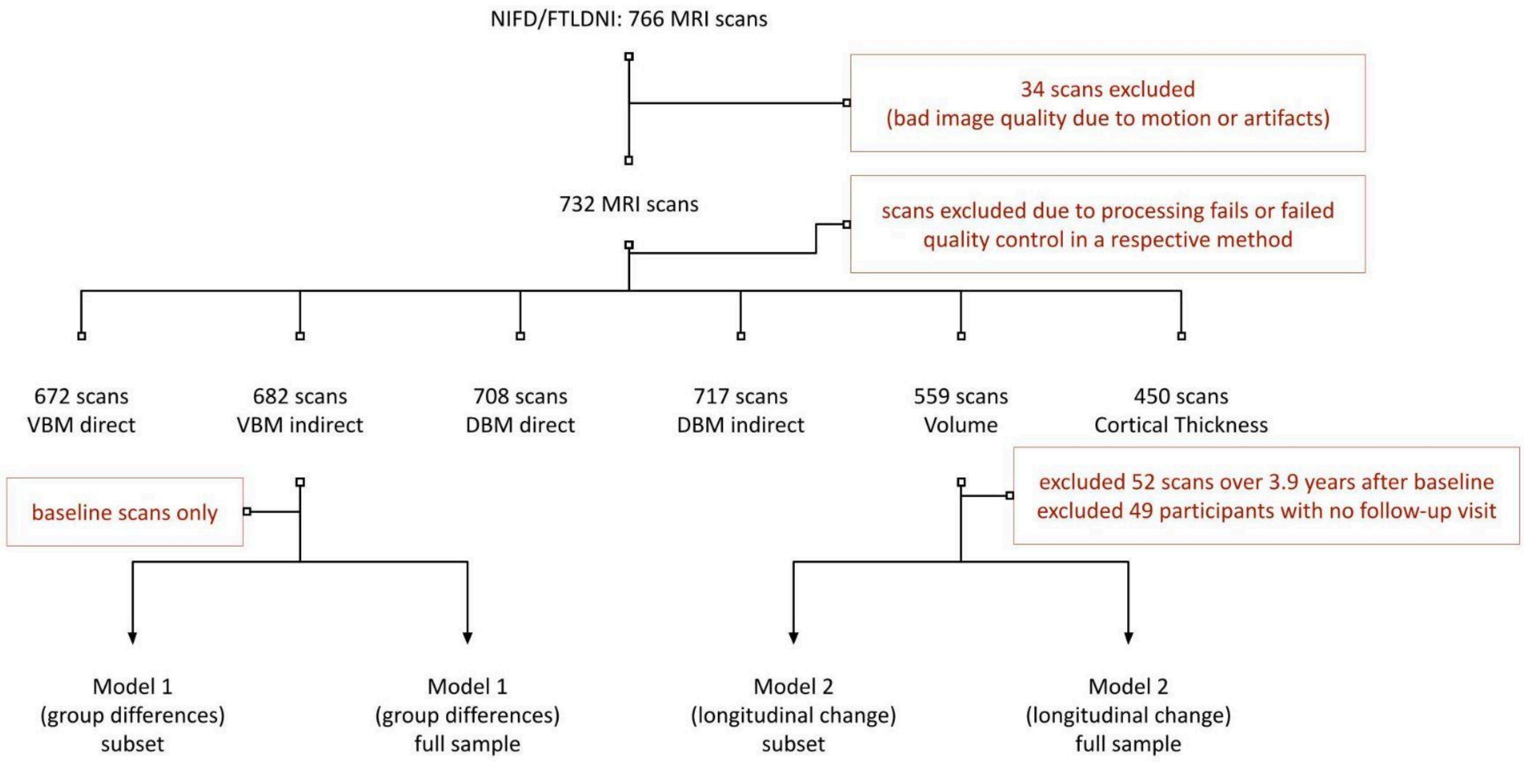


| | Subset | Full sample | | | | | |
|---|---|---|---|---|---|---|---|
| | All methods | Cortical Thickness | Indirect DBM | Direct DBM | Direct VBM | Indirect VBM | FreeSurfer Volumes |
| bvFTD | 21 | 30 | 65 | 53 | 52 | 64 | 43 |
| svPPA | 10 | 18 | 38 | 37 | 37 | 38 | 19 |
| nfvPPA | 15 | 21 | 35 | 34 | 33 | 33 | 28 |
| control | 96 | 108 | 128 | 129 | 127 | 126 | 126 |

| | Subset | Full sample | | | | | |
|---|---|---|---|---|---|---|---|
| | All methods | Cortical Thickness | Indirect DBM | Direct DBM | Direct VBM | Indirect VBM | FreeSurfer Volumes |
| bvFTD | 23 (55) | 32 (79) | 54 (144) | 44 (115) | 44 (114) | 54 (141) | 35 (88) |
| svPPA | 10 (28) | 20 (49) | 37 (103) | 36 (100) | 36 (100) | 37(103) | 18 (49) |
| nfvPPA | 13 (34) | 16 (45) | 30 (82) | 29 (79) | 29 (77) | 30 (79) | 22 (61) |
| control | 88 (214) | 97 (249) | 116 (304) | 117 (306) | 116 (299) | 115 (297) | 112 (295) |

**Flowchart showing the number of participants/scans included in each step of the analyses.** Model 1 table indicates the number of participants per method. Model 2 table shows the number of participants and number of total scans in parentheses.

Supplementary Figure S3: Characteristics of participants whose scans failed PELICAN or FreeSurfer processing

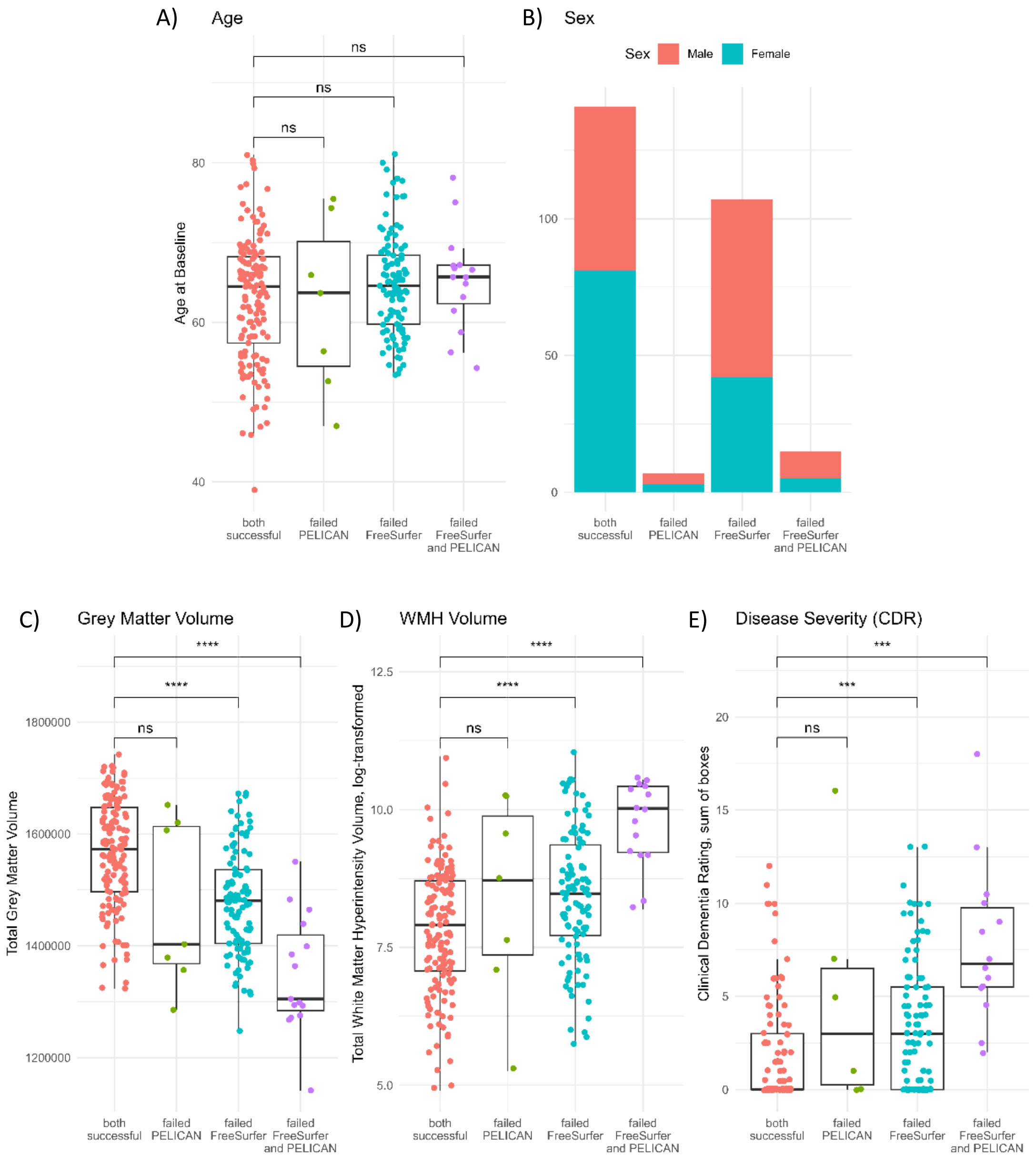


**Baseline demographic and clinical characteristics of participants whose scans failed in either one, both or neither of the tested neuroimaging pipelines FreeSurfer and PELICAN**. Asterisks indicate significant group differences based on one-way ANOVA or χ2 analysis comparing the groups. A) Age. B) Sex. Pipelines failed more often in males. C) Total grey matter volume, based on BISON segmentations. D) Total White Matter Hyperintensity (WMH) volume, based on BISON segmentations of FLAIR images. E) Disease Severity, based on Clinical Dementia Rating (CDR) sum of boxes.

#### Supplementary Figure S4: Additional failed quality control examples for FreeSurfer

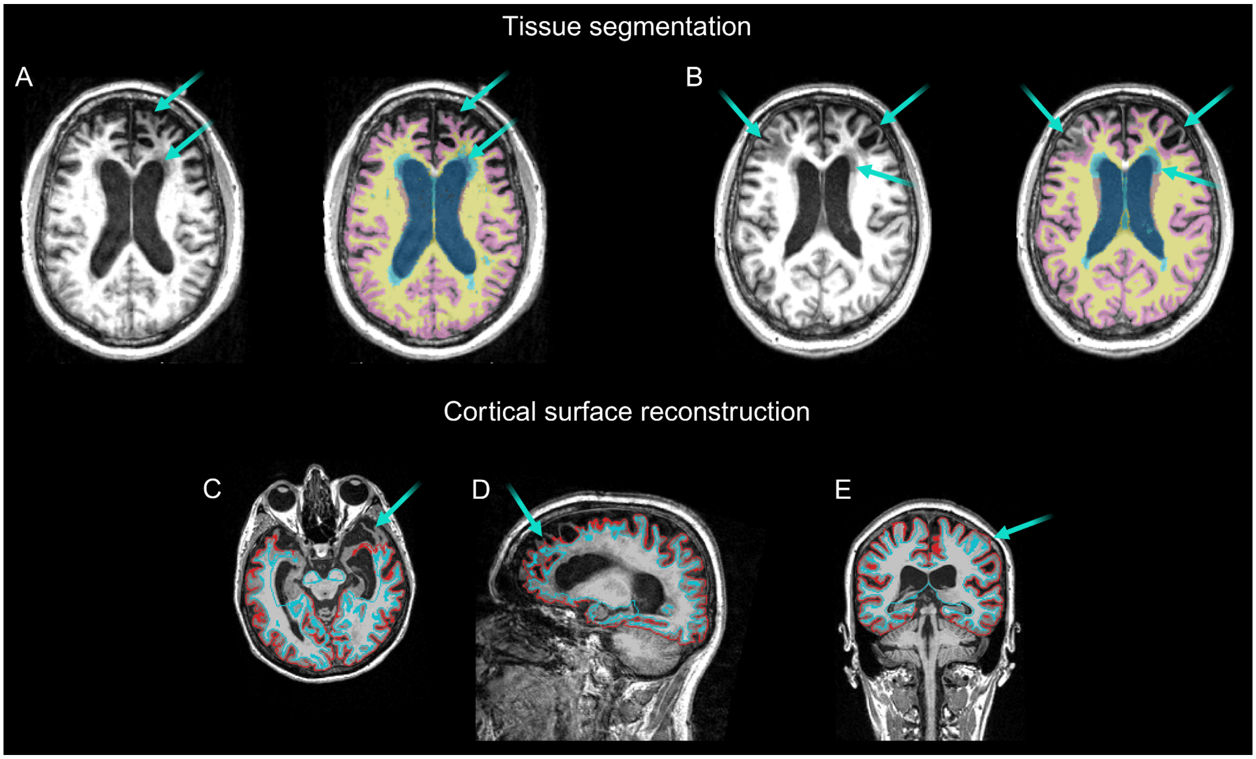


**Examples of failed quality control images for FreeSurfer.** Cyan arrows indicate areas of failure. A, Raw T1w image (left) and tissue segmentation map (right) overlaid on the same image, showing errors in tissue segmentation whereby White Matter Hypointensities were labelled as ventricle and some gyri in the frontal lobe were not identified as grey matter. B, Raw T1w image (left) and tissue segmentation map (right) overlaid on the same image, showing errors in tissue segmentation whereby White Matter Hypointensities were labelled as ventricle and some areas in the frontal lobes, including grey matter and White Matter Hypointensities were not identified as grey matter. C-E, FreeSurfer inner and outer surfaces overlaid on the T1w image in native space. Cortical areas that were either not included in the tesselation (C/D) or parts of th meninges that were erroneously included in as cortical area are indicated with cyan arrows.

#### Supplementary Figure S5: T-value results for linear regression analysis (cross-sectional group differences), subset

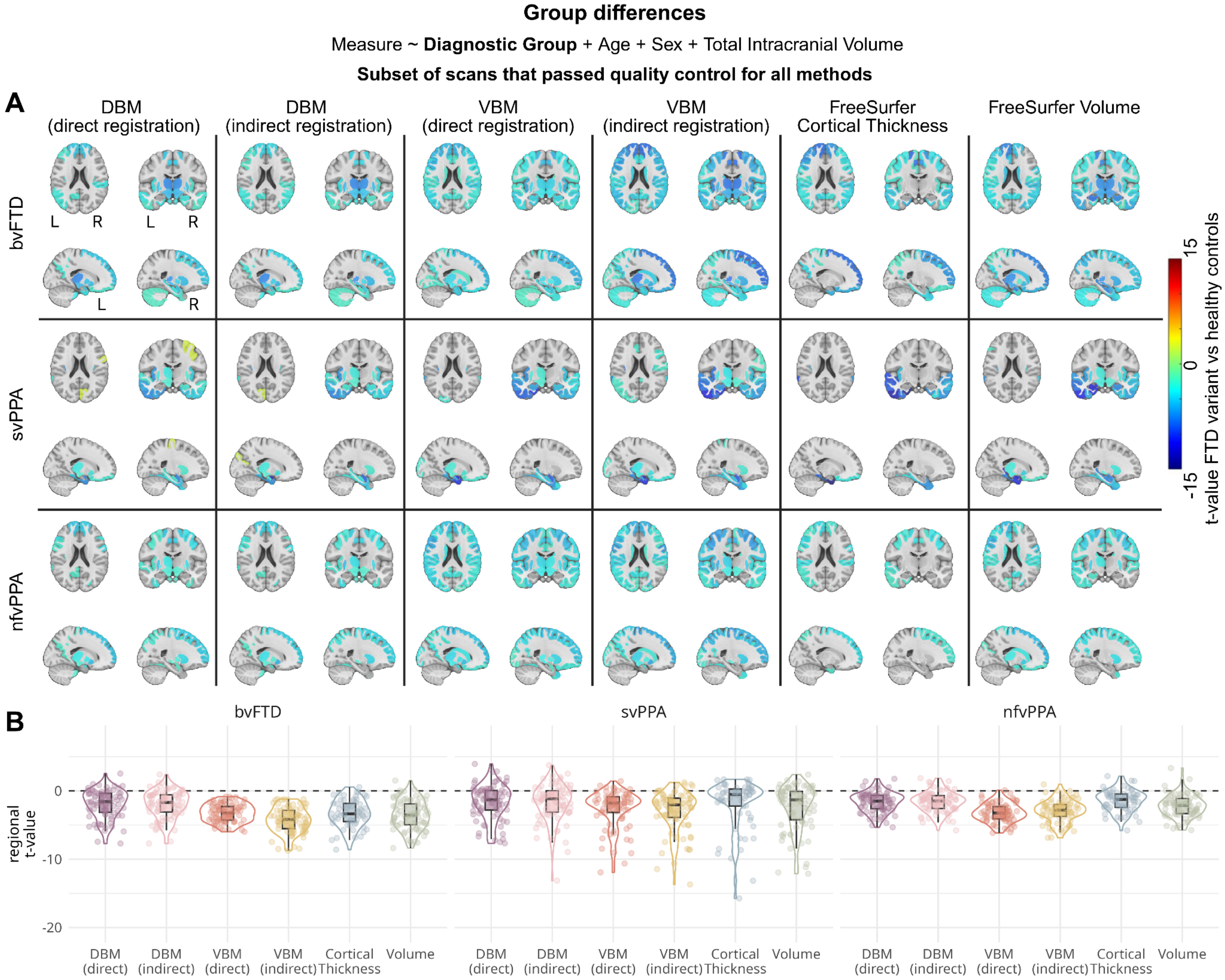


**Results of linear regression models assessing the sensitivity of each method to detect differences between FTD subtypes and healthy controls in the subset of MRI scans that were successfully processed by each method**. A) Brain maps showing t-values for the main effect for diagnostic group, comparing regional values for FTD variants versus healthy controls. B) Box- and violin plots summarizing t-values for the group effect for each method. Each datapoint represents the estimate for one atlas region. VBM: Voxel-Based Morphometry, DBM: Deformation-Based Morphometry, bvFTD: behavioral-variant Frontotemporal Dementia, svPPA: semantic-variant Primary Progressive Aphasia, nfvPPA: nonfluent-variant Primary Progressive Aphasia.

### Supplementary Figure S6: T-value results for linear regression analysis (cross-sectional group differences), full sample

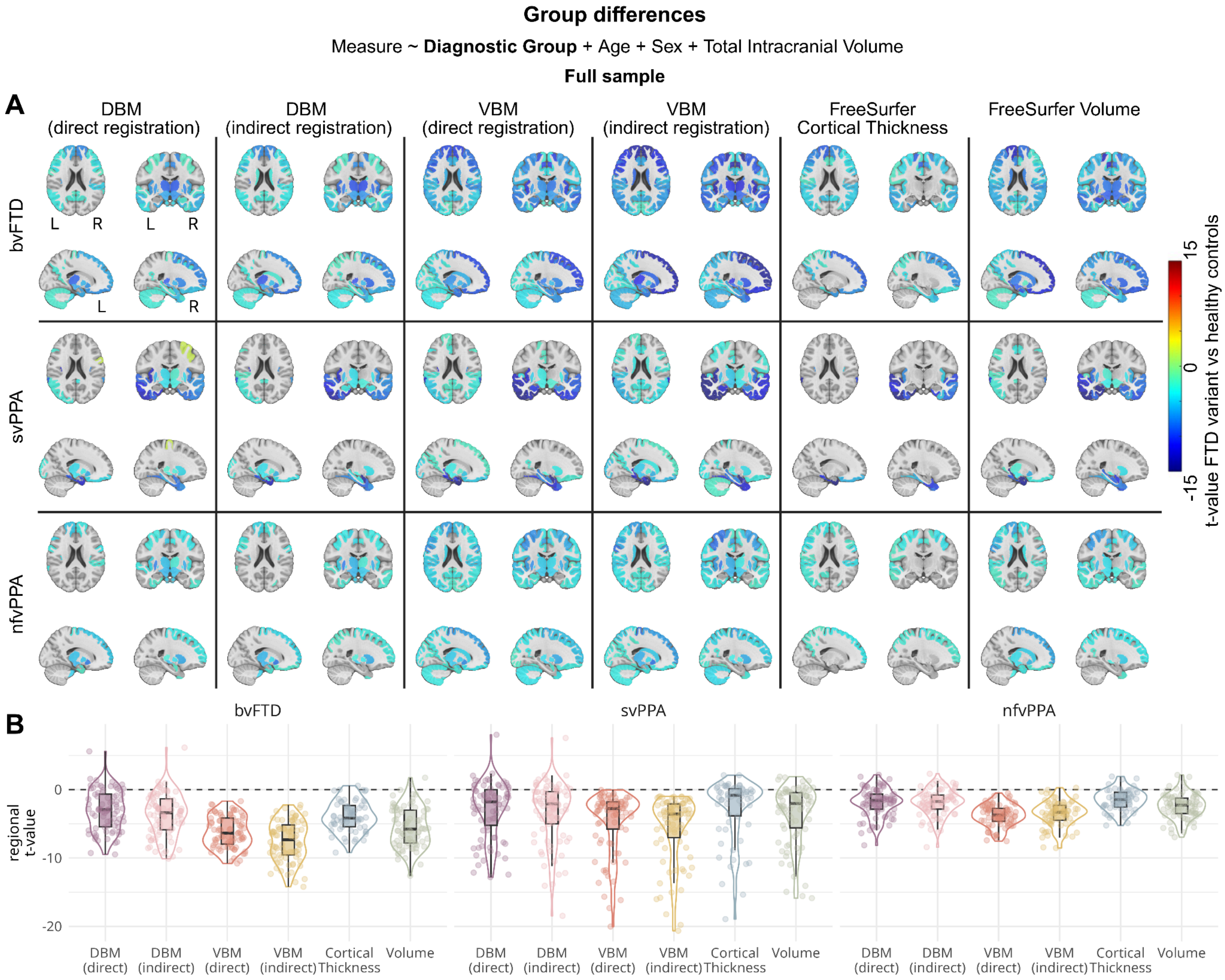


**Results of linear regression models assessing the sensitivity of each method to detect differences between FTD subtypes and healthy controls in the full sample**. A) Brain maps showing t-values for the main effect for diagnostic group, comparing regional values for FTD variants versus healthy controls. B) Box- and violin plots summarizing t-values for the group effect for each method. Each datapoint represents the estimate for one atlas region. VBM: Voxel-Based Morphometry, DBM: Deformation-Based Morphometry, bvFTD: behavioral-variant Frontotemporal Dementia, svPPA: semantic-variant Primary Progressive Aphasia, nfvPPA: nonfluent-variant Primary Progressive Aphasia.

#### Supplementary Figure S7: Beta estimate results for voxel-/vertex-wise linear regression analysis (cross-sectional group differences), subset

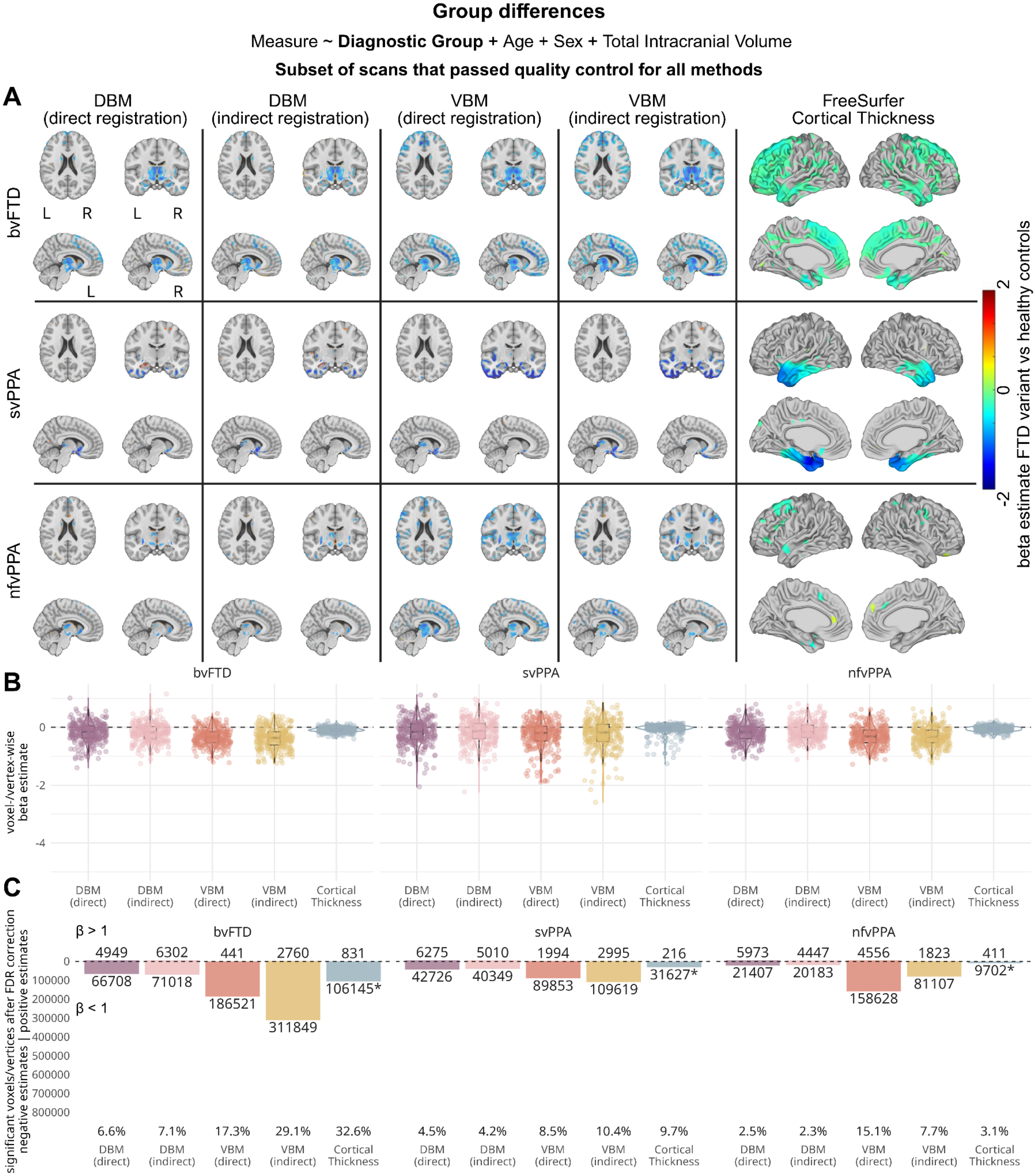

**Results of voxel-/vertex-wise linear regression models assessing the sensitivity of each method to detect differences between FTD subtypes and healthy controls in the subset of MRI scans that were successfully processed by each method**. A) Brain maps showing beta estimates for the main effect for diagnostic group in the grey matter, comparing voxel-/vertex-wise values for FTD variants versus healthy controls. B) Box- and violin plots summarizing beta estimates for the group effect for each method. Each datapoint represents the estimate for one voxel/vertex. For clearer visualization, 300 datapoints were sampled out of the distribution. C) Barplots showing the number of voxels or vertices with significant group differences, after FDR correction, divided by direction of the effect. Percentages of significant voxels or vertices, regardless of effect direction, are shown below each bar. *Note that Cortical Thickness is calculated for 327,684 vertices while VBM and DBM are extracted for 1,082,282 grey matter voxels. VBM: Voxel-Based Morphometry, DBM: Deformation-Based Morphometry, bvFTD: behavioral-variant Frontotemporal Dementia, svPPA: semantic-variant Primary Progressive Aphasia, nfvPPA: nonfluent-variant Primary Progressive Aphasia.

Supplementary Figure S8: T-value results for voxel-/vertex-wise linear regression analysis (cross-sectional group differences), subset

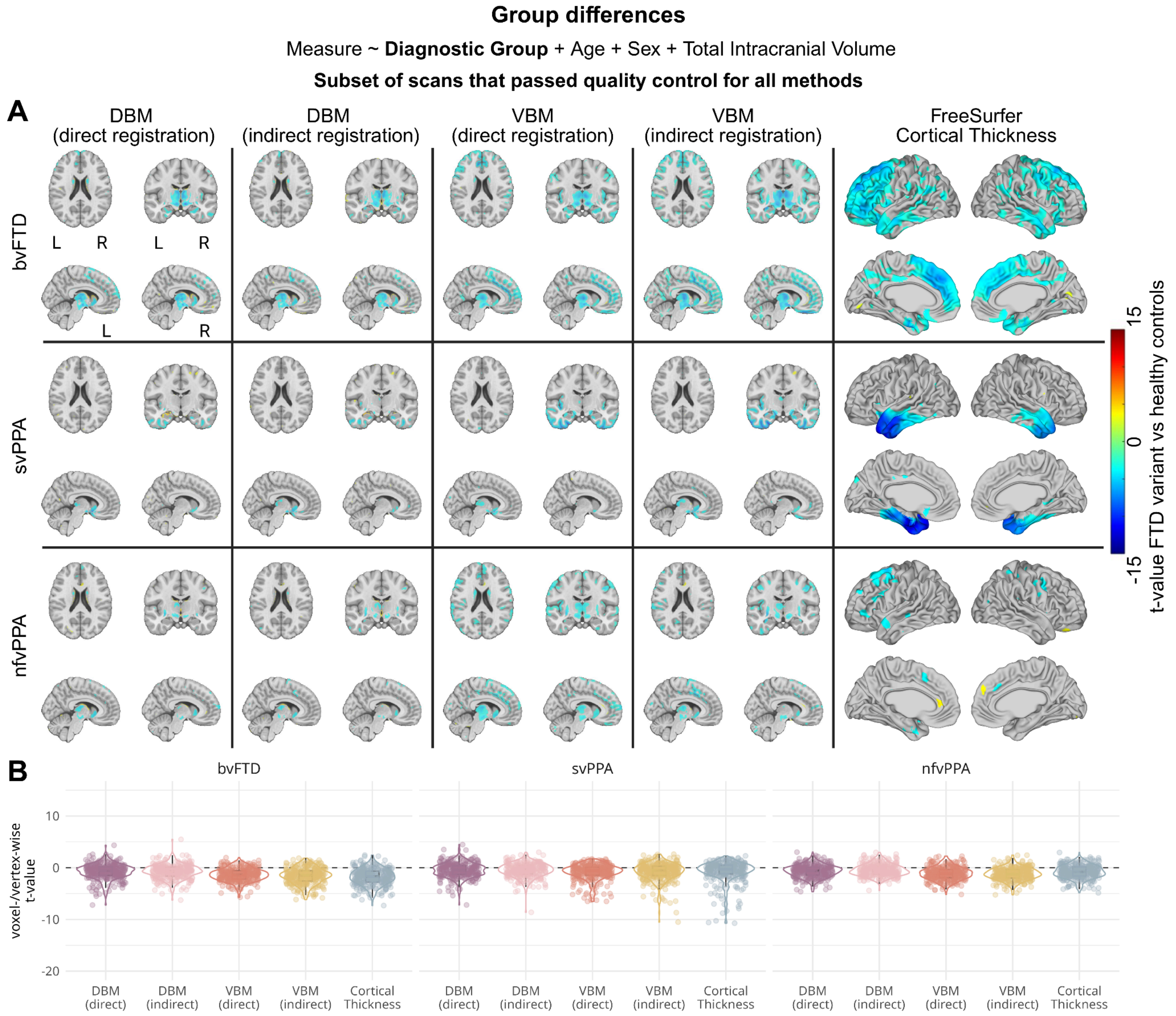


**Results of voxel-/vertex-wise linear regression models assessing the sensitivity of each method to detect differences between FTD subtypes and healthy controls in the subset of MRI scans that were successfully processed by each method**. A) Brain maps showing t-values for the main effect for diagnostic group in the grey matter, comparing voxel-/vertex-wise values for FTD variants versus healthy controls. B) Box- and violin plots summarizing t-values for the group effect for each method. Each datapoint represents the estimate for one voxel/vertex. For clearer visualization, 300 datapoints were sampled out of the distribution. VBM: Voxel-Based Morphometry, DBM: Deformation-Based Morphometry, bvFTD: behavioral-variant Frontotemporal Dementia, svPPA: semantic-variant Primary Progressive Aphasia, nfvPPA: nonfluent-variant Primary Progressive Aphasia.

Supplementary Figure S9: T-value results for voxel-/vertex-wise linear regression analysis (cross-sectional group differences), full sample

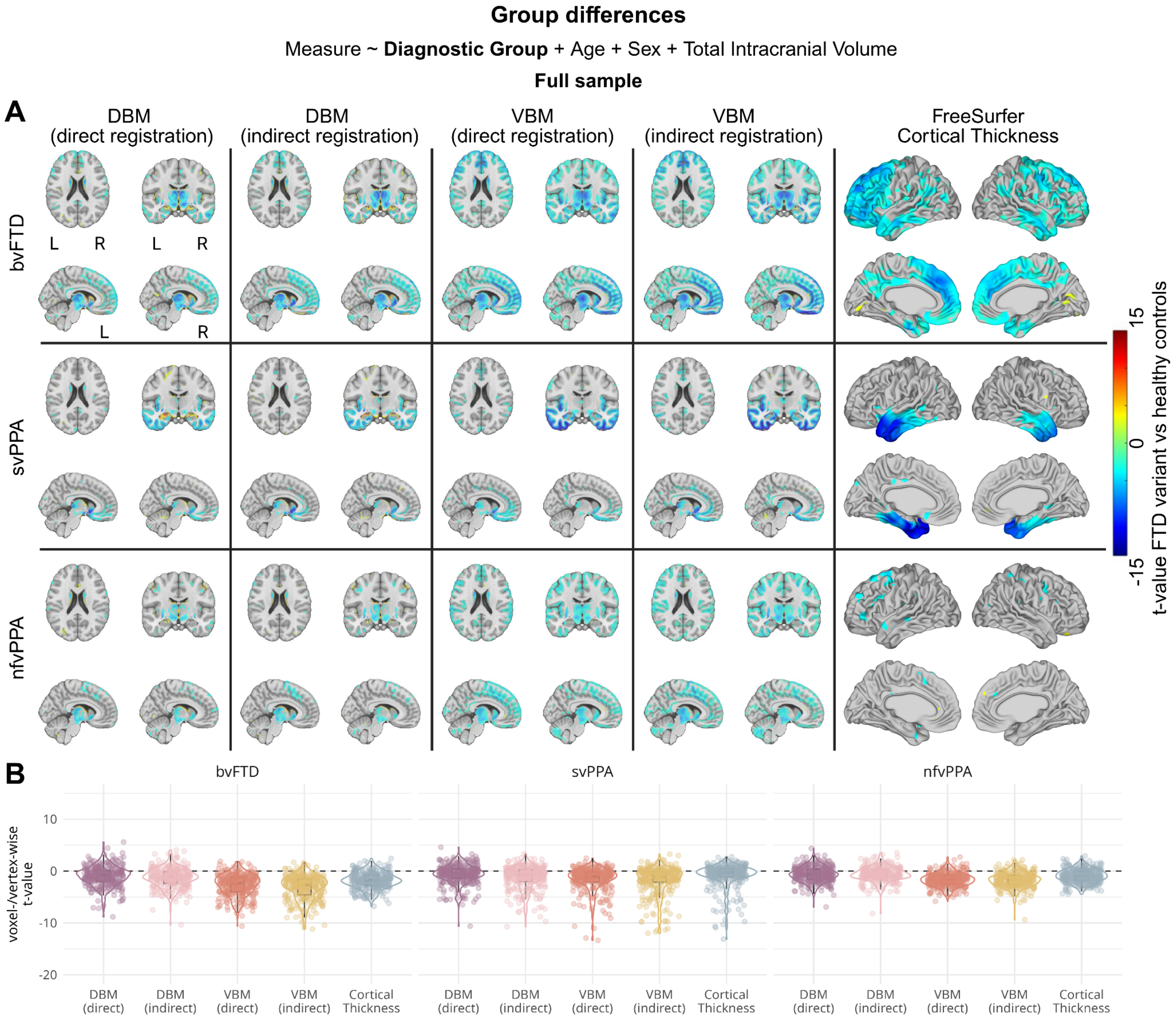


**Results of voxel-/vertex-wise linear regression models assessing the sensitivity of each method to detect differences between FTD subtypes and healthy controls in the full sample**. A) Brain maps showing t-values for the main effect for diagnostic group in the grey matter, comparing voxel-/vertex-wise values for FTD variants versus healthy controls. B) Box- and violin plots summarizing t-values for the group effect for each method. Each datapoint represents the estimate for one voxel/vertex. For clearer visualization, 300 datapoints were sampled out of the distribution. VBM: Voxel-Based Morphometry, DBM: Deformation-Based Morphometry, bvFTD: behavioral-variant Frontotemporal Dementia, svPPA: semantic-variant Primary Progressive Aphasia, nfvPPA: nonfluent-variant Primary Progressive Aphasia.

Supplementary Figure S10: White matter results for voxel-/vertex-wise linear regression analysis (cross-sectional group differences), subset

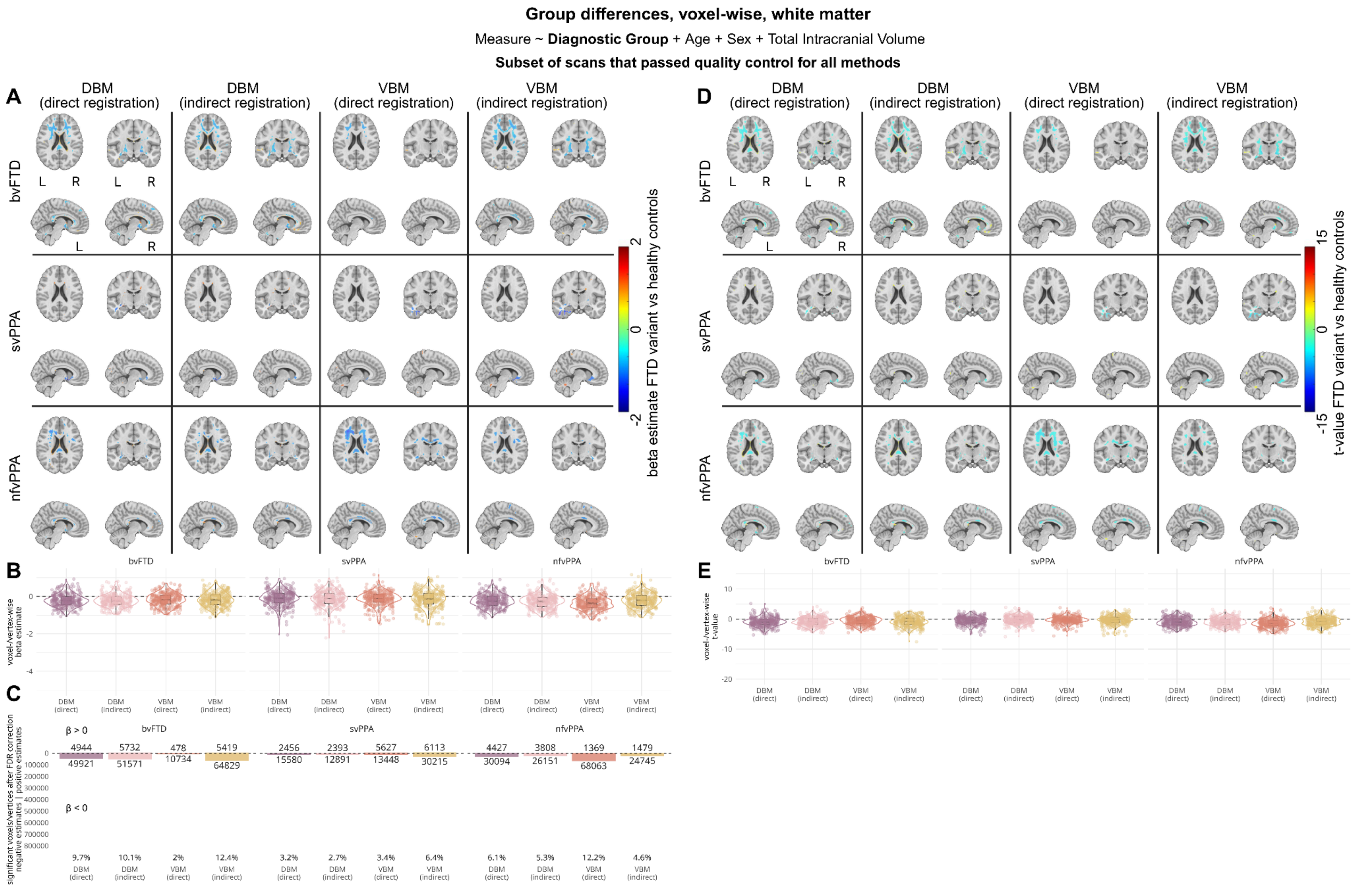

**White matter results of voxel--wise linear regression models assessing the sensitivity of each method to detect differences between FTD subtypes and healthy controls in the subset of MRI scans that were successfully processed by each method**. A) Brain maps showing beta estimates for the main effect for diagnostic group in the grey matter, comparing voxel-wise values for FTD variants versus healthy controls. B) Box- and violin plots summarizing beta estimates for the group effect for each method. Each datapoint represents the estimate for one voxel. For clearer visualization, 300 datapoints were sampled out of the distribution. C) Barplots showing the number of voxels with significant group differences, after FDR correction, divided by direction of the effect. Percentages of significant voxels, regardless of effect direction, are shown below each bar. E) Brain maps showing t-values for the main effect for diagnostic group in the grey matter, comparing voxel-wise values for FTD variants versus healthy controls. F) Box- and violin plots summarizing t-values for the group effect for each method. Each datapoint represents the estimate for one voxel. For clearer visualization, 300 datapoints were sampled out of the distribution. VBM: Voxel-Based Morphometry, DBM: Deformation-Based Morphometry, bvFTD: behavioral-variant Frontotemporal Dementia, svPPA: semantic-variant Primary Progressive Aphasia, nfvPPA: nonfluent-variant Primary Progressive Aphasia.

Supplementary Figure S11: White matter results for voxel-/vertex-wise linear regression analysis (cross-sectional group differences), full sample

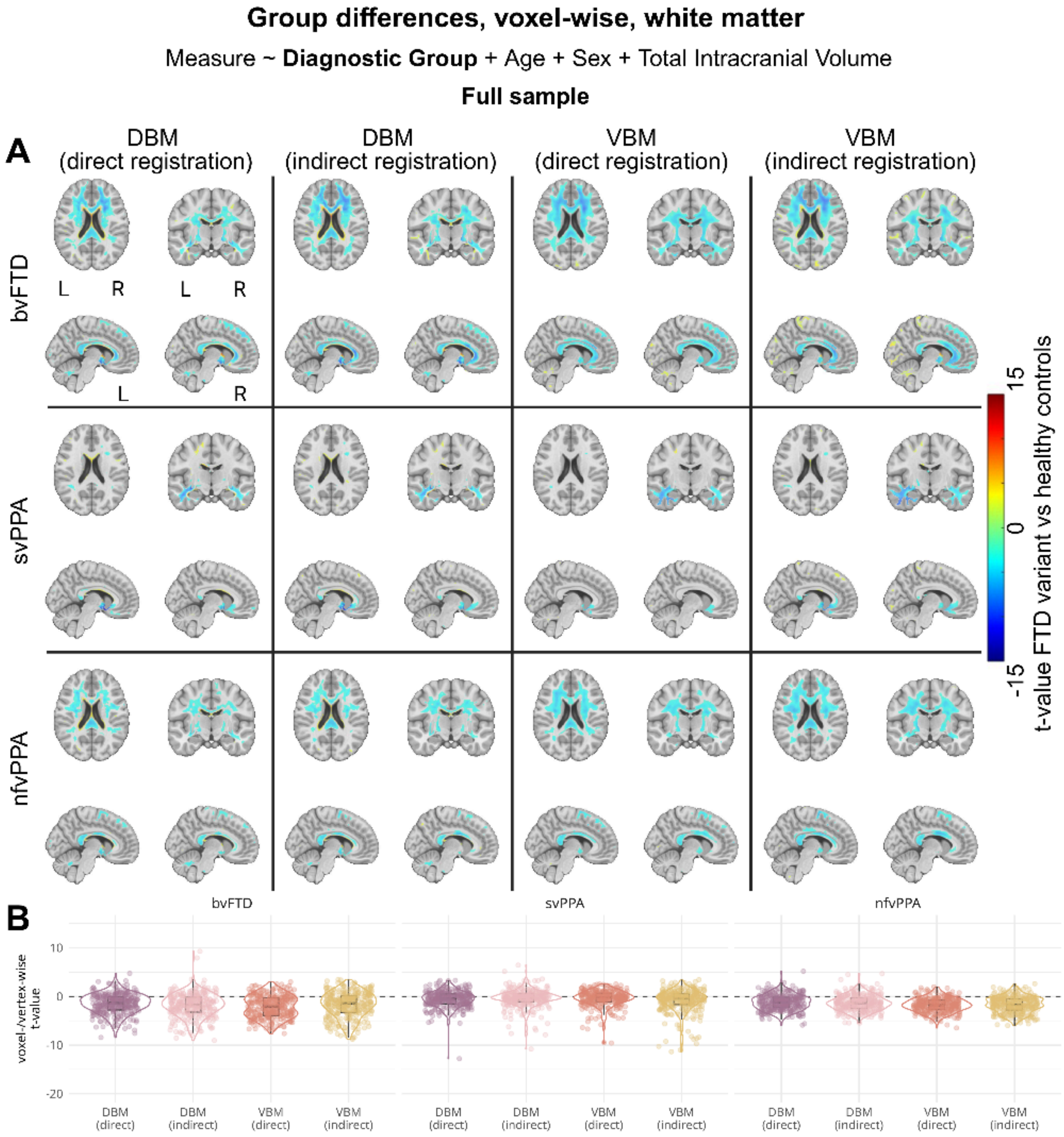


**White matter results of voxel--wise linear regression models assessing the sensitivity of each method to detect differences between FTD subtypes and healthy controls in the full sample**. A) Brain maps showing beta estimates for the main effect for diagnostic group in the grey matter, comparing voxel-wise values for FTD variants versus healthy controls. B) Box- and violin plots summarizing beta estimates for the group effect for each method. Each datapoint represents the estimate for one voxel. For clearer visualization, 300 datapoints were sampled out of the distribution. C) Barplots showing the number of voxels with significant group differences, after FDR correction, divided by direction of the effect.

Percentages of significant voxels, regardless of effect direction, are shown below each bar. E) Brain maps showing t-values for the main effect for diagnostic group in the grey matter, comparing voxel-wise values for FTD variants versus healthy controls. F) Box- and violin plots summarizing t-values for the group effect for each method. Each datapoint represents the estimate for one voxel. For clearer visualization, 300 datapoints were sampled out of the distribution. VBM: Voxel-Based Morphometry, DBM: Deformation-Based Morphometry, bvFTD: behavioral-variant Frontotemporal Dementia, svPPA: semantic-variant Primary Progressive Aphasia, nfvPPA: nonfluent-variant Primary Progressive Aphasia.

### Supplementary Figure S12: T-value results for linear mixed-effects analysis (longitudinal change), subset

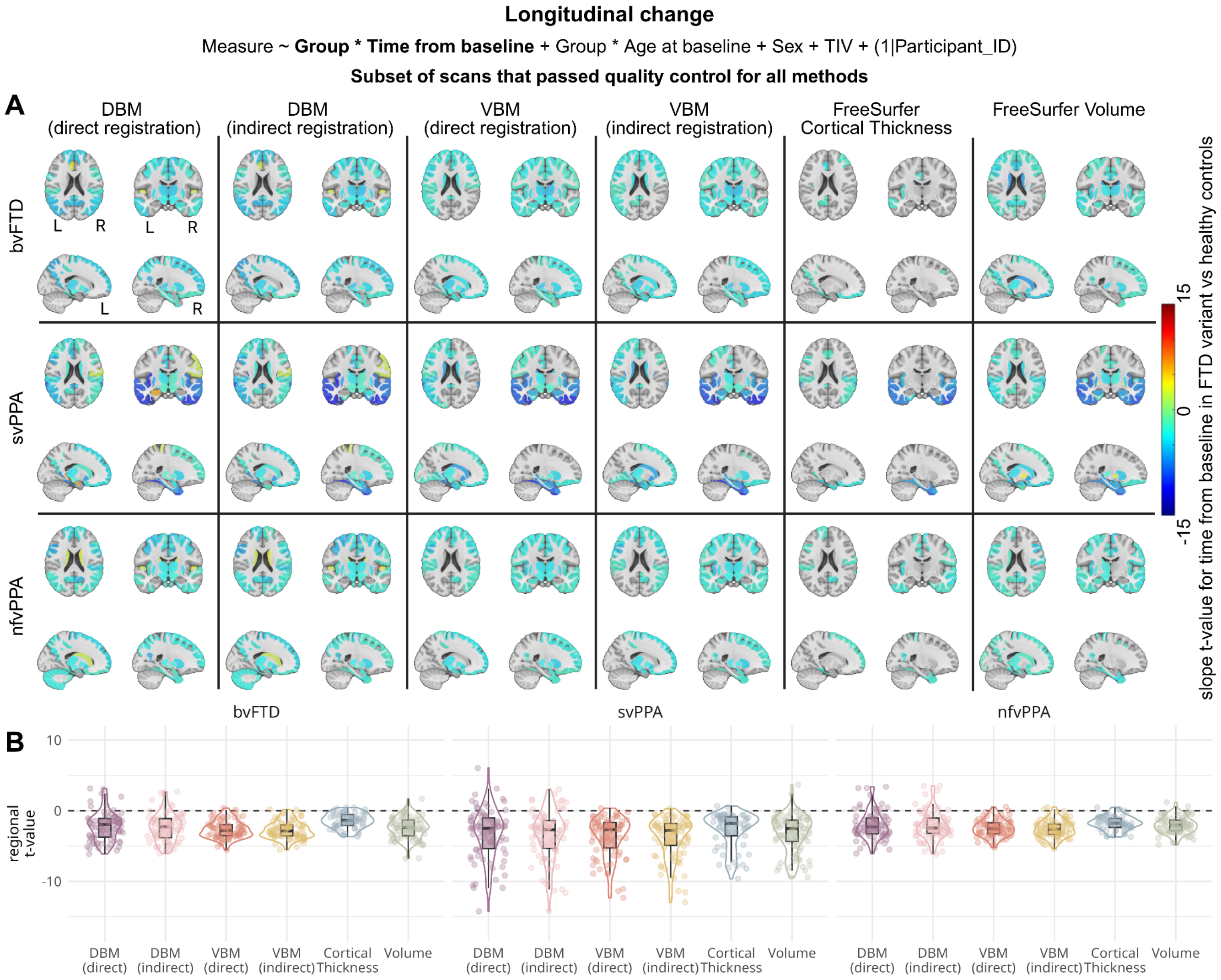


**Results of linear mixed-effects models assessing the sensitivity of each method to detect longitudinal changes between FTD subtypes and healthy controls in the subset of MRI scans that were successfully processed by each method**. A) Brain maps showing t-values for the interaction between diagnostic group and time from baseline, comparing regional slopes for FTD variants versus healthy controls. B) Box- and violin plots summarizing t-values for the interaction effect for each method. Each datapoint represents the estimate for one atlas region. VBM: Voxel-Based Morphometry, DBM: Deformation-Based Morphometry, bvFTD: behavioral-variant Frontotemporal Dementia, svPPA: semantic-variant Primary Progressive Aphasia, nfvPPA: nonfluent-variant Primary Progressive Aphasia.

#### Supplementary Figure S13: T-value results for linear mixed-effects analysis (longitudinal change), full sample

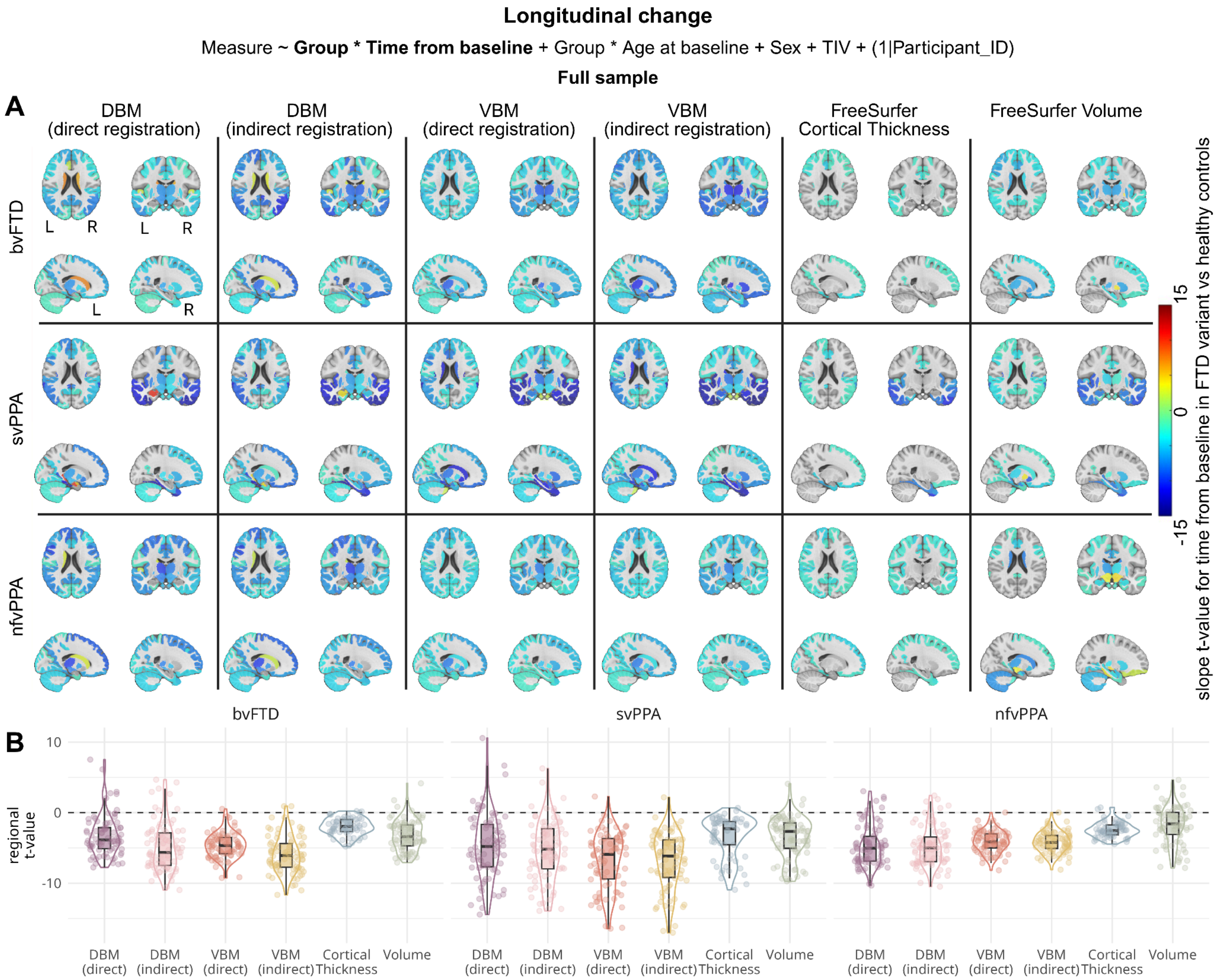


**Results of linear mixed-effects models assessing the sensitivity of each method to detect longitudinal changes between FTD subtypes and healthy controls in the full sample**. A) Brain maps showing t-values for the interaction between diagnostic group and time from baseline, comparing regional slopes for FTD variants versus healthy controls. B) Box- and violin plots summarizing t-values for the interaction effect for each method. Each datapoint represents the estimate for one atlas region. VBM: Voxel-Based Morphometry, DBM: Deformation-Based Morphometry, bvFTD: behavioral-variant Frontotemporal Dementia, svPPA: semantic-variant Primary Progressive Aphasia, nfvPPA: nonfluent-variant Primary Progressive Aphasia.

### Supplementary Figure S14: CAT12 output examples

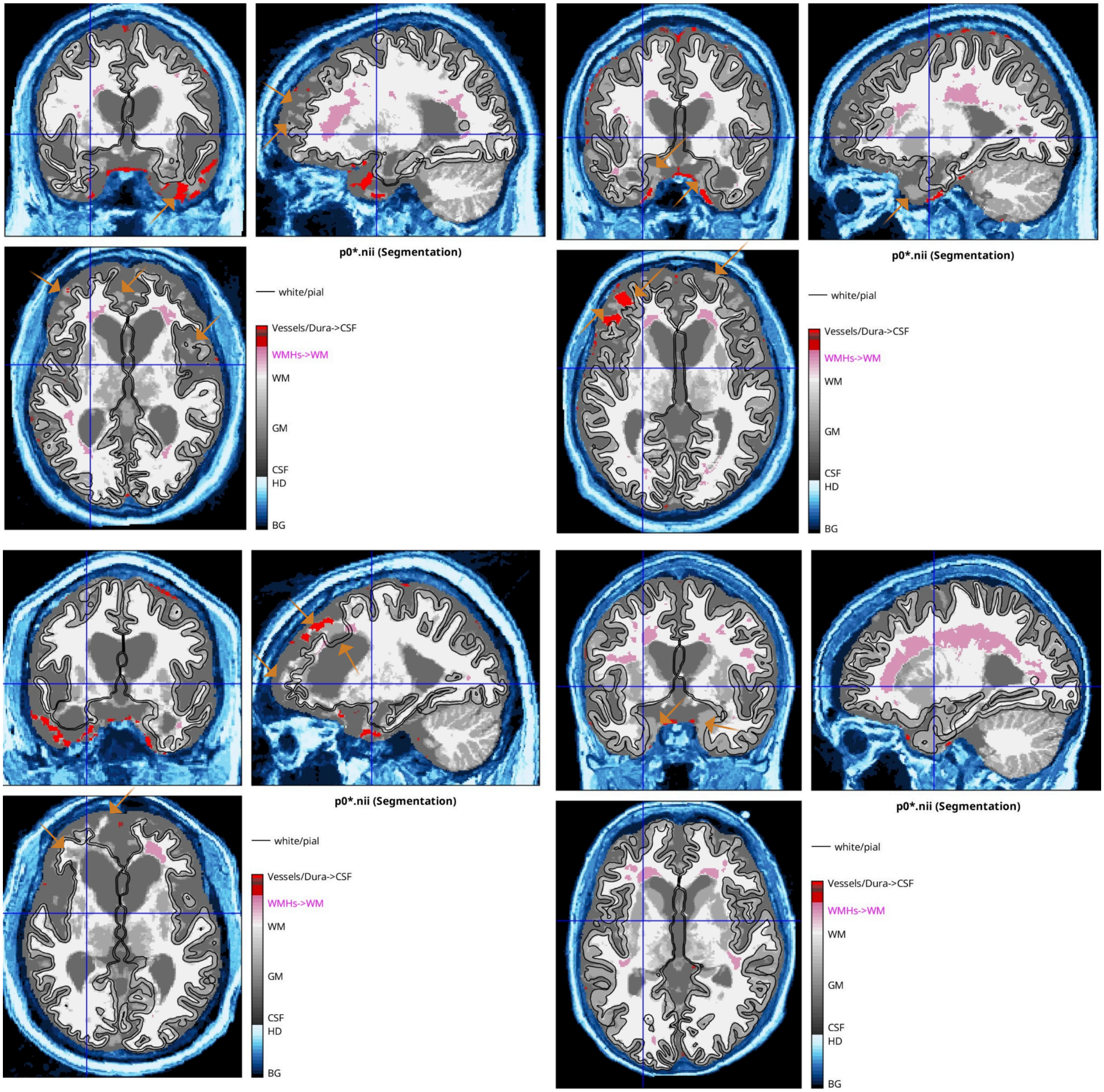


**Example of a failed quality control image for CAT12 tissue segmentation mask overlaid on the linearly registered T1w image.** Major failures include not capturing frontal and temporal lobe cortical regions in areas of severe atrophy, as indicated by orange arrows.
